# Brain imaging and neuropsychological assessment of individuals recovered from a mild to moderate SARS-CoV-2 infection

**DOI:** 10.1101/2022.07.08.22277420

**Authors:** Marvin Petersen, Felix Leonard Nägele, Carola Mayer, Maximilian Schell, Elina Petersen, Simone Kühn, Jürgen Gallinat, Jens Fiehler, Ofer Pasternak, Jakob Matschke, Markus Glatzel, Raphael Twerenbold, Christian Gerloff, Götz Thomalla, Bastian Cheng

## Abstract

As SARS-CoV-2 infections have been shown to affect the central nervous system, the investigation of associated alterations of brain structure and neuropsychological sequelae is crucial to help address future health care needs. Therefore, we performed a comprehensive neuroimaging and neuropsychological assessment of 223 non-vaccinated individuals recovered from a mild to moderate SARS-CoV-2 infection (100 female/123 male, age [years], mean ± SD, 55.54 ± 7.07; median 9.7 months after infection) in comparison with 223 matched controls (93 female/130 male, 55.74 ± 6.60) within the framework of the Hamburg City Health Study. Primary study outcomes were advanced diffusion magnetic resonance imaging (MRI) measures of white matter microstructure, cortical thickness, white matter hyperintensity load and neuropsychological test scores. Among all 11 MRI markers tested, significant differences were found in global measures of mean diffusivity and extracellular free-water which were elevated in the white matter of post-SARS-CoV-2 individuals comparing to matched controls (free-water: 0.148 ± 0.018 vs. 0.142 ± 0.017, *P*<.001; mean diffusivity [10^−3^ mm^2^/s]: 0.747 ± 0.021 vs. 0.740 ± 0.020, *P*<.001). Group classification accuracy based on diffusion imaging markers was up to 80%. Neuropsychological test scores did not significantly differ between groups. Collectively, our findings suggest that subtle changes in white matter extracellular water content last beyond the acute infection with SARS-CoV-2. However, in our sample, a mild to moderate SARS-CoV-2 infection was not associated with neuropsychological deficits, significant changes in cortical structure or vascular lesions several months after recovery. External validation of our findings and longitudinal follow-up investigations are needed.

**Significance statement:** In this case-control study, we demonstrate that non-vaccinated individuals recovered from a mild to moderate SARS-CoV-2 infection show significant alterations of the cerebral white matter identified by diffusion weighted imaging, such as global increases in extracellular free-water and mean diffusivity. Despite the observed brain white matter alterations in this sample, a mild to moderate SARS-CoV-2 infection was not associated with worse cognitive functions within the first year after recovery. Collectively, our findings indicate the presence of a prolonged neuroinflammatory response to the initial viral infection. Further longitudinal research is necessary to elucidate the link between brain alterations and clinical features of post-SARS-CoV-2 individuals.

## Introduction

As the number of patients recovering from an acute infection with the severe acute respiratory syndrome coronavirus type 2 (SARS-CoV-2) grows, the study of its long-term consequences on health outcomes has gained much attention (1–4).

It is widely recognized that coronavirus disease 2019 (COVID-19) caused by SARS-CoV-2 not only leads to respiratory dysfunction, but also impacts various other organ systems during the acute phase and well beyond (1, 5, 6). Neurological symptoms, such as headache, fatigue, memory and attention deficits, may significantly impede well-being in individuals suffering from COVID-19 sequelae (4, 7, 8). Advancing our understanding of the underlying pathological mechanisms will be crucial for addressing future health care needs.

Different potential mechanisms have been suggested to be involved in the development and persistence of neurological symptoms in patients with COVID-19. Post-mortem histopathological and molecular studies have demonstrated viral neurotropism, signs of neuroinflammation (9, 10), neurodegeneration (11), demyelination (12), axonal disruption (13), as well as micro- and macrovascular damage (14, 15). However, most studies were conducted in patients with severe COVID-19, whereas histopathological findings from individuals with mild to moderate courses are lacking.

*In vivo* studies applying modern brain imaging joined by comprehensive clinical and neuropsychological assessment are scarce. Recent preliminary evidence from the UK Biobank suggests cortical thickness reductions in the olfactory and limbic network, as well as neurocognitive decline in former COVID-19 patients, although these findings still need to be replicated in independent datasets (11). The majority of remaining studies focused on visually apparent pathological findings such as intracranial hemorrhage, stroke or white matter hyperintensities in small case series or single case reports of more severely affected patients (16–19). Taken together, current evidence is of limited transferability to patients with a mild to moderate SARS-CoV-2 infection, therefore necessitating further investigations.

In order to address this research need, we studied 223 non-vaccinated individuals in median 289 days after recovery from mainly mild to moderate SARS-CoV-2 infections in a retrospective, cross-sectional case-control design. We leveraged advanced magnetic resonance imaging (MRI) techniques enabling the study of imaging phenotypes associated with neurodegeneration, atrophy, myelin/cellular disruption, inflammation, as well as vascular damage (20–23). Moreover, study participants received a comprehensive clinical and neuropsychological assessment. Building upon our previous multi-organ assessment in this cohort (1), here, we provide a detailed *in vivo* assessment of the cerebral white and gray matter, as well as neuropsychological outcomes in former COVID-19 patients.

## Results

### Sample characteristics

We examined participants of the Hamburg City Health Study (HCHS) and its COVID Program. Imaging data was available for 230 post-SARS-CoV-2 individuals. Following quality assessment (QA), in total 7 post-SARS-CoV-2 individuals had to be excluded, leaving 223 cases for propensity-score matching with healthy controls who had passed QA. Results of the matching procedure are visualized in **Figure S1**. The final sample included 223 matched controls (93 female, age in years, mean ±standard deviation [SD], 55.74 ± 6.60) and 223 post-SARS-CoV-2 individuals (100 female, 55.54 ± 7.07, see **Table 1**). Of the latter, the majority had a mild to moderate course of COVID-19 (without symptoms, n=7; mild symptoms, n=125; moderate symptoms, n=67), 18 were hospitalized and none required mechanical ventilation or intensive care unit treatment. There were no significant differences between post-SARS-CoV-2 individuals and matched controls regarding age, sex, years of education and cardiovascular risk factors.

**Table 1.** Baseline characteristics of post-SARS-CoV-2 individuals and matched controls.

|  | Post-SARS-CoV-2<br>individuals (N = 223) | Matched controls<br>(N = 223) | *P<sub>uncorr</sub>* |
| --- | --- | --- | --- |
| Demographics |  |  |  |
| Age in years, mean ± SD | 55.54 ± 7.07 | 55.74 ± 6.60 | .76 |
| Female sex, N (%) | 100 (44.8) | 93 (41.7) | .56 |
| Education in years, mean ±<br>SD | 15.70 ± 2.56 | 15.67 ± 2.86 | .91 |
| COVID-19-specific characteristics |  |  |  |
| Self-reported disease severity at the time of infection |  |  |  |
| Asymptomatic, N (%) | 7 (3.1) |  |  |
| Mild, N (%) | 125 (56.1) |  |  |
| Moderate, non-hospitalized, N (%) | 67 (30.0) |  |  |
| Moderate, hospitalized,<br>without ICT, N (%) | 18 (8.1) |  |  |
| Days between first positive<br>SARS-CoV-2 PCR test and<br>study enrollment, median<br>(IQR) | 289 (163, 318) |  |  |
| Cardiovascular risk factors |  |  |  |
| Hypertension<sup>a</sup>, N (%) | 131 (58.7) | 121 (54.3) | .39 |
| Dyslipidemia<sup>b</sup>, N (%) | 54 (24.2) | 51 (22.9) | .82 |
| Diabetes mellitus<sup>c</sup>, N (%) | 16 (7.2) | 13 (5.8) | .70 |
| Smoking, ever, N (%) | 107 (48.0) | 105 (47.1) | .92 |

*Abbreviations:* COVID-19 = coronavirus disease 2019, ICT = intensive-care treatment, IQR = inter-quartile range, PCR = polymerase chain reaction, post-SARS-CoV-2 individuals = individuals who recovered from a severe acute respiratory coronavirus type 2 infection, SD = standard deviation
<sup>a</sup>Prevalence of hypertension was defined as blood pressure $\geq 140/90$ mmHg, intake of antihypertensive medication or self-report
<sup>b</sup>Prevalence of dyslipidemia was defined as LDL-cholesterol/HDL-cholesterol ratio $> 3.5$ or intake of lipid lowering therapies
<sup>c</sup>Prevalence of diabetes mellitus was defined as fasting blood glucose level $>126$ mg/dl or self-report

### Clinical data

Although post-SARS-CoV-2 individuals showed nominally better test performances in Verbal Fluency (VF), Mini Mental State Examination (MMSE), and clock drawing test (CDT) compared to matched controls, after Bonferroni-correction for multiple comparisons, no significant group differences remained in any neuropsychological test score, including those associated with executive functioning, memory, psychosocial and neurological symptom burden (**Table 2**).

**Table 2.** Results of clinical and neuropsychological assessments of post-SARS-CoV-2 individuals compared to matched controls.

| Clinical measure<sup>a</sup> | Post-SARS-CoV-2 individuals | Matched controls | $P_{uncorr}$<br>b | $P_{bonf}$<sup>c</sup> | $F$ |
| --- | --- | --- | --- | --- | --- |
| Neurocognition |  |  |  |  |  |
| TMT-A in seconds | 31.89 ± 10.60 (212) | 33.71 ± 11.67 (190) | .12 | >.99 | 2.40 |
| TMT-B in seconds | 68.50 ± 22.69 (212) | 70.89 ± 25.57 (187) | .37 | >.99 | .81 |
| VF | 28.03 ± 6.04 (212) | 26.43 ± 7.15 (212) | .02 | .14 | 5.94 |
| WLR | 8.52 ± 1.63 (210) | 8.32 ± 1.61 (204) | .25 | >.99 | 1.33 |
| MMSE | 28.37 ± 1.26 (211) | 28.02 ± 1.72 (210) | .02 | .19 | 5.34 |
| CDT | 6.75 ± 0.78 (212) | 6.57 ± 1.03 (214) | .04 | .37 | 4.20 |
| Psychosocial symptom burden |  |  |  |  |  |
| PHQ-9 | 3.94 ± 3.74 (212) | 3.91 ± 3.77 (215) | .97 | >.99 | <.01 |
| GAD-7 | 2.94 ± 3.28 (212) | 2.80 ± 3.06 (215) | .67 | >.99 | .18 |
| Neurological symptom burden |  |  |  |  |  |
| PHQ-15<sup>d</sup> | 2.13 ± 1.83 (212) | 1.83 ± 1.73 (215) | .09 | .82 | 2.86 |

*Abbreviations:* CDT = clock drawing test, GAD = General Anxiety Disorder, MMSE = Mini Mental State Examination, PHQ = Patient Health Questionnaire, post-SARS-CoV-2
individuals = individuals who recovered from a severe acute respiratory coronavirus type 2 infection, SD = standard deviation, TMT-A = Trail-Making-Test Part A, TMT-B = TMT Part B, VF = verbal fluency, WLR = word list recall
<sup>a</sup>Presented as mean ± SD (N)
<sup>b</sup>Uncorrected *P* values of analyses of covariance, adjusted for age, sex and years of education
<sup>c</sup>Bonferroni-corrected *P* values of analyses of covariance, adjusted for age, sex and years of education (considering 9 comparisons)
<sup>d</sup>PHQ-15 items: headache, dizziness, fatigue, sleep disturbances

### Imaging

We first conducted analyses of covariance, adjusted for sex, age and education, to test for group differences in imaging markers averaged across the entire white and gray matter. Post-SARS-CoV-2 individuals exhibited higher global extracellular free-water and mean diffusivity (MD) relative to matched controls, markers associated with immune activation and atrophy (mean ± SD, free-water: 0.148 ± 0.018 vs. 0.142 ± 0.017, *F*=18.47, *P*_*bonf*_<.001; MD [10^−3^ mm^2^/s]: 0.747 ± 0.021 vs. 0.740 ± 0.020, *F*=17.28, *P*_*bonf*_<0.001) (**Figure 2, Table S1**). While peak width of skeletonized mean diffusivity (PSMD) (*P*_*uncorr*_=.005), a marker of cerebral small vessel disease, and cortical thickness (*P*_*uncorr*_=.01) were nominally increased in post-SARS-CoV-2 individuals, both measures, as well as the remaining averaged imaging markers of white matter fiber structure were not significantly different between groups after Bonferroni-correction (**Figure 2, Table S1**).

**Figure 1.**
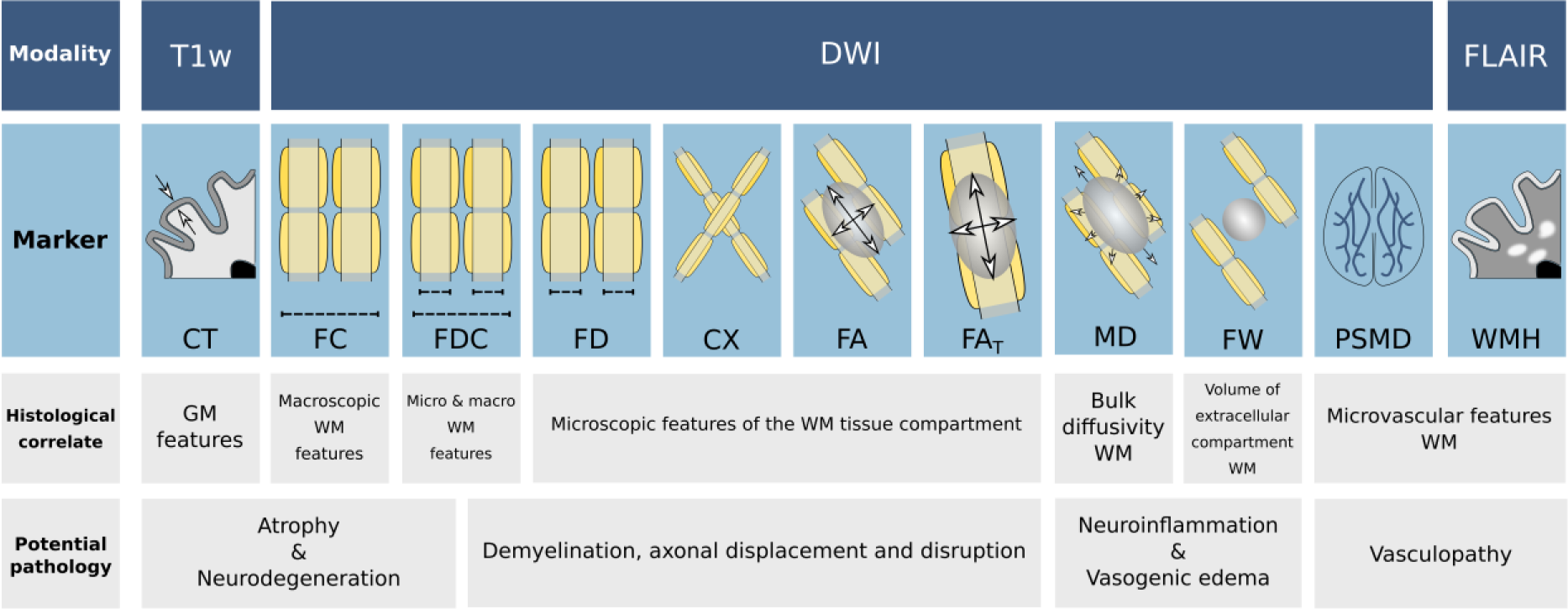
Schematic illustration of the investigated imaging markers. To assess the cerebral gray and white matter, micro- and macrostructural imaging markers were derived. The first row of the schematic describes the imaging sequences utilized to derive the imaging markers below. The second row presents diagrammatic illustrations of the markers: CT was determined as the distance between the pial surface and white matter/gray matter boundary; FC represents the macroscopic white matter fiber-bundle diameter; FD reflects the microscopic intraaxonal volume; as the combinatorial measure of FC and FD, FDC simultaneously assesses micro- and macroscopic alterations of white matter tracts; CX measures the intricacy of fiber configurations within a voxel; FA measures the directional preference of diffusion; MD denotes the molecular diffusion rate; free-water imaging enables the adjustment of traditional diffusion tensor imaging markers for extracellular diffusion signal, which increases their tissue specificity (FA_T_); FW represent the volume of the extracellular compartment; PSMD was calculated as the difference of the 95^th^ and 5^th^ percentile of skeletonized MD values; WMH load represents the white matter hyperintensity volume normalized by the total intracranial volume. Histological interpretations of the respective imaging markers and their potential sensitivity for pathologies are described in the third and fourth row, respectively. *Abbreviations*: CT = cortical thickness, CX = complexity, FA = fractional anisotropy, FA_T_ = FA of the tissue, FD = fiber density, FDC = fiber density and cross-section, FLAIR = fluid-attenuated inversion recovery, FW = free-water, Log. FC = logarithm of fiber cross-section, MD = mean diffusivity, PSMD = peak width of skeletonized MD, WMH = white matter hyperintensity.

**Figure 2.**
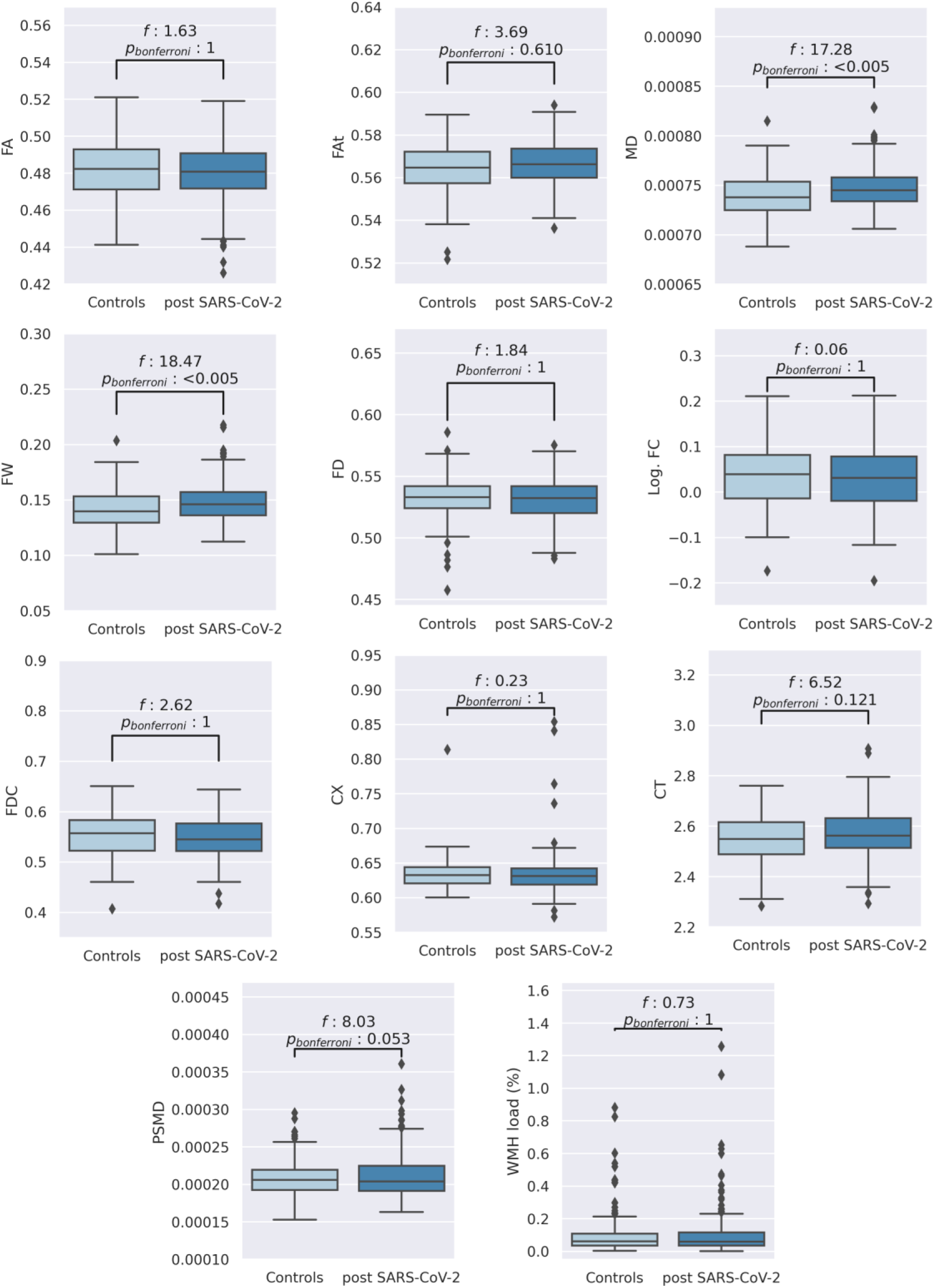
Group comparison of neuroimaging indices on a global scale. Boxplots of averaged imaging measures and the corresponding statistical results (F-statistics and Bonferroni-corrected *P* values) from the ANCOVAs comparing matched controls with post-SARS-CoV-2 individuals adjusted for age, sex, and years of education. Abbreviations: CT = cortical thickness, CX = complexity, FA = fractional anisotropy, FA_T_ = FA of the tissue, FD = fiber density, FDC = fiber density and cross-section, FW = free-water, Log. FC = logarithm of fiber cross-section, MD = mean diffusivity, post-SARS-CoV-2 = individuals who recovered from a severe acute respiratory coronavirus type 2 infection, PSMD = peak width of skeletonized MD, WMH = white matter hyperintensity

To detect spatial patterns of brain structural alterations, we additionally performed vertex- and voxel-wise analyses of gray and white matter imaging markers. Vertex-wise comparisons of cortical thickness did not reveal significant differences between matched controls and post-SARS-CoV-2 participants. Voxel-wise statistics on the entire white-matter skeleton, a representation of major white matter fiber bundles, revealed predominant free-water and MD increases in the white matter skeleton of post-SARS-CoV-2 subjects encompassing all brain lobes, comparing to very localized changes in other diffusion markers (**Figure 3, Table S2**). More specifically, the conventional diffusion tensor imaging markers fractional anisotropy (FA) and MD showed significant differences between groups, with FA increases in 0.8% and decreases in 1.2% of the skeleton in cases relative to healthy controls. MD was significantly increased in 41.3% and decreased in 0.1% of the skeleton of post-SARS-CoV-2 participants. Employing free-water imaging, post-SARS-CoV-2 individuals showed significant free-water elevations in 38.3% and reductions in 0.4% of the skeleton, as well as FA of the tissue (FA_T_) elevations in 3.3% of the skeleton, but no FA_T_ reductions. Alterations of the remaining diffusion markers were of even less spatial extent (<3%).

**Figure 3.**
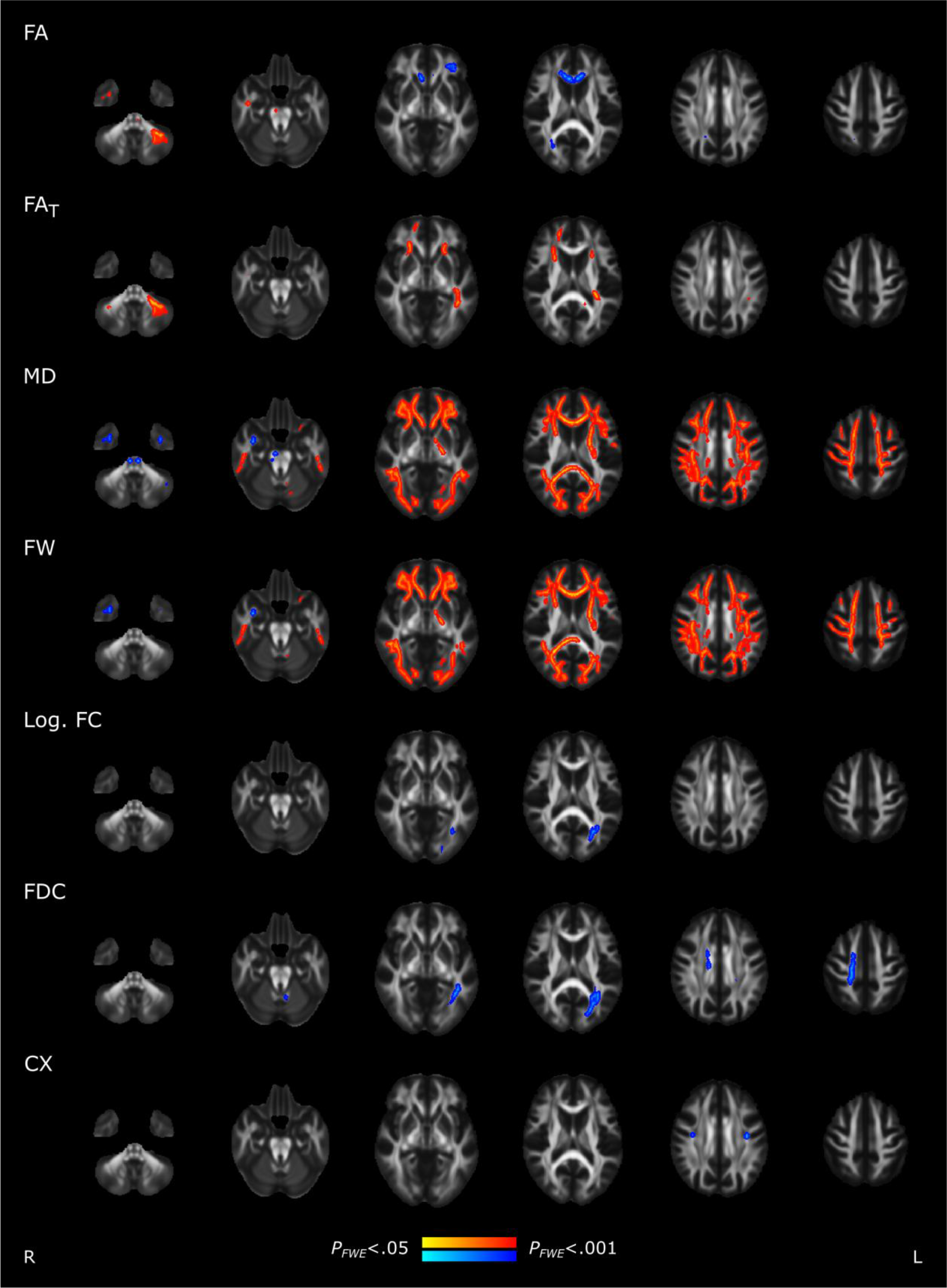
Group comparison of skeletonized diffusion indices. Skeleton voxels that significantly differed between groups are highlighted by colors: post-SARS-CoV-2 individuals > matched controls, red; post-SARS-CoV-2 individuals < matched controls, blue. Abbreviations: CX = complexity, FA = fractional anisotropy, FA_T_ = FA of the tissue, FD = fiber density, FDC = fiber density and cross-section, FW = free-water, FWE = family-wise error corrected, Log. FC = logarithm of fiber cross-section, MD = mean diffusivity, post-SARS-CoV-2 = individuals who recovered from a severe acute respiratory coronavirus type 2 infection

### Associations between clinical and imaging data

Exploratory regression analyses were performed between clinical measures and averaged imaging markers that showed significant group differences, i.e., free-water and MD.

Linear regression revealed a significant positive association of free-water with Trail-Making-Test Part A (TMT-A) (*P*=.008) and Part B (TMT-B) (*P*<.001), as well as significant negative associations of free-water with VF (*P*=.003), and Word List Recall (WLR) (*P*<.001) in the entire sample (**Table S3**). MMSE, CDT, Patient Health Questionnaire-9 (PHQ-9), General Anxiety Disorder-7 (GAD-7) and PHQ-15 scores were not significantly correlated with free-water. Moreover, we observed significant group x free-water interactions for VF (*P*=.006), WLR (*P*=.02), MMSE (*P*=.02) and CDT (*P*=.04). Post-hoc Spearman correlations performed for matched controls and post-SARS-CoV-2 individuals, separately, confirmed positive associations with TMT-A (*rho*=0.20, *P*=.004) and TMT-B (*rho*=0.22, *P*=.001), as well as negative correlations with VF (*rho*=-0.23, *P*<.001) and WLR (*rho*=-0.25, *P*<.001) in the post-SARS-CoV-2 group. However, among all neuropsychological measures; free-water was only significantly correlated with TMT-B (*rho*=0.15, *P*=.04) in the group of matched controls (**Table S3, Figure S2**).

Results for MD were very similar. Linear regression analyses showed significant positive associations of MD with TMT-A (*P* =.03) and TMT-B (*P*=.001), as well as negative associations of MD with VF (*P*=.01) and WLR (*P*<.001) in the combined group of matched controls and post-SARS-CoV-2 individuals. Further, significant group x MD interactions were present for TMT-A, VF, WLR, MMSE and CDT (**Table S4**). Post-hoc Spearman correlations revealed significant positive correlations of MD with TMT-A (*rho*=0.17, *P*=.01) and TMT-B (*rho*=0.20, *P*=.005), as well as negative correlations of MD with VF (*rho*=-0.22, *P*=.001) and WLR (*rho*=-0.23, *P*<.001) in the post-SARS-CoV-2 group only. All other correlations were non-significant (**Table S4, Figure S3**).

### Prediction of a past SARS-CoV-2 infection based on imaging markers

We examined the predictive capacity of derived imaging markers employing a supervised machine learning approach (**Figure 4**). Free-water (80.21%) and MD (79.38%) achieved the strongest median prediction accuracies. The median cortical thickness score was 45.95%. All investigated metrics but cortical thickness scored significantly better than null models for which the group assignment was randomly permuted.

**Figure 4.**
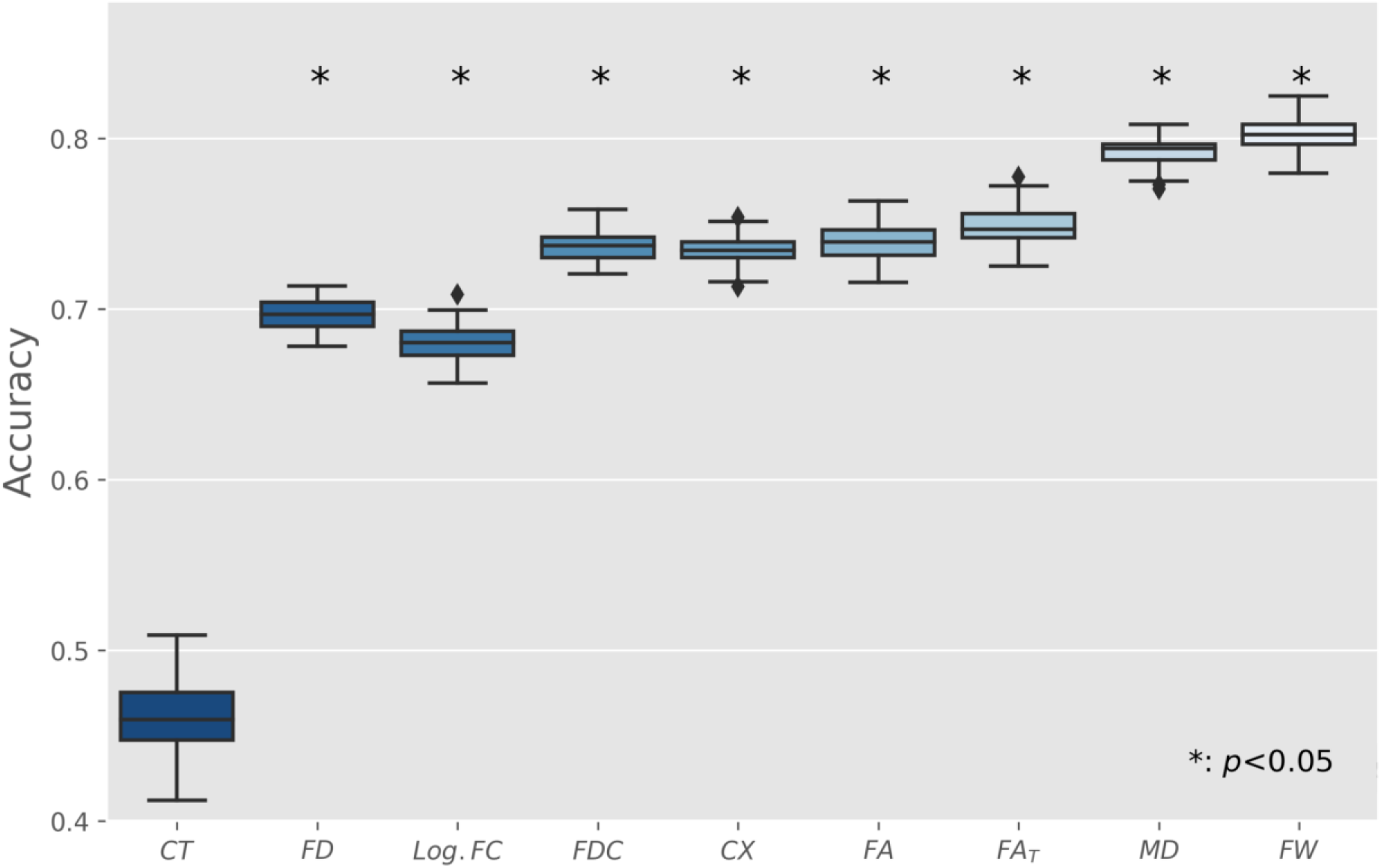
Prediction of past SARS-CoV-2 infection based on imaging markers. Boxplots represent the accuracy of models trained in a 10-fold nested cross-validation setup. To address scoring being biased by a single arbitrary split of training and test sets, predictions have been repeated 100 times for each marker with different random split regimens. Asterisks indicate significant difference to null-model predictions. *Abbreviations*: CT = cortical thickness, CX = complexity, FA = fractional anisotropy, FA_T_ = FA of the tissue, FD = fiber density, FDC = fiber density and cross-section, FW = free-water, Log. FC = logarithm of fiber cross-section, MD = mean diffusivity, PSMD = peak width of skeletonized MD, WMH = white matter hyperintensity

### Sensitivity analyses

Analysis results remained stable if formerly hospitalized post-SARS-CoV-2 individuals were excluded and if post-SARS-CoV-2 individuals were stratified by recruitment route (see **SI Appendix**).

## Discussion

We investigated brain structural alterations and neuropsychological sequelae in a large sample of individuals who recovered from mainly mild to moderate COVID-19. At median 289 days after the acute infection, these individuals showed significantly higher average free-water and MD in the white matter compared to matched healthy controls. In contrast, cortical thickness and markers of cerebral small vessel disease were not significantly different between groups. In addition, white matter diffusion indices successfully predicted a past SARS-CoV-2 infection. We did not detect neuropsychological deficits post-SARS-CoV-2 individuals. Collectively, our study suggests that a mild to moderate SARS-CoV-2 infection is associated with subtle microstructural alterations in the cerebral white matter beyond the stage of acute infection.

A key aspect of COVID-19 neuropathology appears to be the dynamic response of the intrathecal immune system to the virus. Evidence of neuroinflammation was reported in histopathological and clinical studies of COVID-19 patients: virus invasion (24, 25), activation of glial cells (9, 26, 27), and a cytokine response in the cerebrospinal fluid accompanying neurological and psychiatric COVID-19 symptoms (10). In our study, we observed widespread increases of extracellular free-water and MD in post-SARS-CoV-2 individuals encompassing all brain lobes. Both free-water and MD are sensitive to an activated immune response causing excessive extracellular free-water and thus increased diffusivity (28–30). More specifically, microglia and astrocytes emit cytokines upon activation, inducing osmosis of water from the blood into the extracellular space (31, 32). Interestingly, endothelial dysfunction and subsequent vascular leakage due to persistent immune activation has been previously implicated in the pathophysiology of COVID-19 (33, 34). Taken together, it is conceivable that the observed increase in free-water and MD could be an indirect sign of a prolonged neuroinflammatory reaction to a SARS-CoV-2 infection. Nevertheless, other possible mechanisms for changes in the extracellular space need to be considered.

Volume increases in the extracellular compartment might be accompanied by structural damage like demyelination as well as axonal disruption secondary to neuroinflammation. Free-water corrected diffusion markers enable further guidance in microstructural interpretations. While analyses of overall mean value showed no significant group differences in free-water corrected FA_T_, voxel-wise statistics identified increased FA_T_ in corresponding frontal areas of post-SARS-CoV-2 individuals. Yet, these changes included only ∼3% of the white matter skeleton, indicating either subtle or spatially limited effects localized to association tracts. Normal to increased FA_T_ in the presence of elevated free-water suggests minor microstructural alterations like axonal compression or displacement rather than damage to myelin sheaths or axons which would rather lead to FA_T_ decreases (21). Moreover, fixel markers, which also model properties of the tissue compartment (35, 36), did not show group differences averaged across the entire white matter skeleton. Thus, in contrast to previous work demonstrating more widespread FA reductions in small samples of hospitalized COVID-19 cases (37–39), our findings suggest that white matter changes following a mild to moderate SARS-CoV-2 infection most likely reflect subtle increases in extracellular free-water as opposed to structural neural damage.

Based on previous histopathological reports of vasculopathy in COVID-19 and higher ACE-2 expression in cells of the blood-brain-barrier, we hypothesized that post-SARS-CoV-2 individuals would show alterations in imaging markers of small vessel disease burden (14). However, in our study, WMH load was not significantly different, indicating that a mild to moderate course of COVID-19 does not lead to visually accessible vascular lesions (WMH) as previously reported (40). PSMD, an established imaging marker of small vessel disease more sensitive to microstructural changes (23), showed nominally higher values in post-SARS-CoV-2 individuals, but differences did not survive Bonferroni-correction. Taken together, follow-up investigations are needed to understand whether subtle long-term vascular impairments will eventually increase the prevalence of cerebrovascular disease among COVID-19 convalescents (33).

Alterations of cortical gray and white matter commonly co-occur in neurological diseases (41, 42). Notably, this was not the case in our study. This is contrasted by a recent report on mildly-affected COVID-19 subjects in the UK Biobank demonstrating longitudinal volumetric reductions in the gray matter in olfactory networks (11). On the other hand, a current cross-sectional study has shown gray matter volume increases in long-COVID patients compared to healthy controls (43). The discrepancies between these studies and our work may be due to general differences in recruitment strategies (general population vs enriched samples of individuals suffering from long-term sequelae) and study designs (longitudinal vs. cross-sectional), both of which likely affect the sensitivity to detect gray matter changes associated with a SARS-CoV-2 infection.

By providing scores of prediction accuracy, our machine learning analysis evaluated brain imaging markers for their diagnostic relevance. Logistic regression models based on free-water and MD achieved a considerable accuracy of ∼80% in predicting a past SARS-CoV-2 infection, outperforming other imaging markers under study. Of note, cortical thickness achieved the lowest accuracy, not significantly differing from prediction by chance. The difference in accuracy between diffusion metrics and cortical thickness might highlight that, in mild to moderate COVID-19, pathophysiological aspects are better detected by diffusion imaging-based techniques. Finally, the higher accuracy of fiber cross-section compared to cortical thickness – both morphometric measures – might imply that COVID-19-associated alterations preferably occur in the white matter.

We want to emphasize, that our results represent average effects, i.e., not every mild to moderate affected COVID-19 patient may exhibit the reported changes. In addition, our results are based on a non-vaccinated cohort. As vaccination has been repetitively demonstrated to be a highly effective measure against COVID-19, vaccinated patients possibly exhibit less of the pathophysiological substrates identified in our study (44).

It is important to put the observed brain white matter alterations into a clinical perspective. Reported persisting clinical sequelae of COVID-19 include executive dysfunction, anxiety, depression, fatigue, muscle weakness and sleep impairment (11, 45– 47). In contrast, we found no significant difference for any cognitive domain, depression, anxiety and neurological symptoms between groups. Besides the absence or mild expression of the respective symptoms, other reasons, such as the relatively long follow-up period in our investigation, as well as differing degrees of social deprivation as a result of country-specific pandemic control measures, may explain the discrepancy with other studies. Exploration of imaging-behavior interactions showed that relatively increased free-water and MD were associated with worse executive performance (TMT-A/B), working memory (WLR) and verbal fluency (VF) in post-SARS-CoV-2 individuals, thus providing a preliminary pathophysiological link between neurocognitive deficits and brain alterations in individuals recovered from COVID-19. Clearly, more research is needed to increase our understanding of factors underlying the persistence of neurological symptoms in a sub-group of COVID-19 patients.

Strengths of this work lie in its considerable sample size; high quality imaging and phenotypical data; a robust and reproducible image processing pipeline; the investigation happening at an early stage of the pandemic, potentially alleviating the problem of different COVID-19 strains and vaccinations as confounders; and a conservative statistical correction scheme to reduce the false-positive rate.

However, our study also exhibits limitations. Our investigation lacks information about SARS-CoV-2 strains as well as precise disease severity stratification beyond the self-reported information on hospitalization and subjective perception of disease intensity. In addition, we follow a cross-sectional study design unable to fully address premorbid group differences despite a rigorous matching procedure. Future longitudinal studies could not only elaborate on the trajectory of the identified microstructural white matter alterations, but also address the question whether these findings are markers of an increased susceptibility for the development of neurological sequelae.

We performed a comprehensive assessment of established neuroimaging markers for structural neural integrity to characterize neurobiological changes potentially underlying post-acute COVID-19 neuropsychological sequelae after a mainly mild to moderate disease course. Our findings support the notion of a prolonged neuroinflammatory response indicated by subtle, but widespread increases in extracellular free-water and mean diffusivity in the white matter of COVID-19 convalescents. In contrast, we did not observe signs of cortical atrophy or macrostructural vascular damage. Importantly, despite identifying this characteristic imaging footprint, the investigated sample exhibited no marked neuropsychological symptoms 10 months after SARS-CoV-2 infection. External validation and longitudinal investigations are needed to further clarify the clinical relevance of our findings.

## Materials and methods

### Study population

We examined participants of the Hamburg City Health Study (HCHS) COVID Program. A detailed description of the study design has been published previously (1). Post-SARS-CoV-2 participants [1] had a positive polymerase chain reaction (PCR) test for SARS-CoV-2 and [2] were aged between 45 and 74 at inclusion. Recruitment routes included both invitation upon laboratory-confirmed SARS-CoV-2 infection and self-referral of participants following newspaper announcement. Subsequent to recruitment, the participants underwent the study protocol of the HCHS (48) – including cranial MR imaging, neuropsychological testing and a self-report questionnaire on COVID-19-associated symptoms. In addition, a healthy control group was sampled from the original HCHS cohort (48). The previously reported matching procedure (1) considered confounders allowing for a comprehensive investigation of COVID-19 pathophysiology in multiple organ systems beyond the brain, including the lungs, heart, vasculature and kidneys. We refined this procedure to specifically account for confounds known to affect cognitive performance as well as indices derived from structural and diffusion MR imaging: the groups were matched for age, sex, years of education as well as for the prevalence of arterial hypertension, diabetes mellitus, dyslipidemia and smoking behavior by propensity scores using the matchit (v4.3.3) R package (49).

### Ethics approval

The local ethics committee of the Landesärztekammer Hamburg (State of Hamburg Chamber of Medical Practitioners, PV5131) approved the study and written informed consent was obtained from all participants (1, 50).

### Clinical assessments

Cognitive testing was performed by a trained study nurse and included the MMSE (51), TMT-A/B (52), VF and WLR subtests of the Consortium to Establish a Registry for Alzheimer’s Disease Neuropsychological Assessment Battery (CERAD-Plus) (53), as well as the CDT (54). Psychosocial symptom burden was evaluated by the GAD-7 (anxiety) (55) and PHQ-9 (depression) (56). Moreover, self-reported neurological symptoms (headache, dizziness, fatigue, sleep disturbances) were assessed by part of the PHQ-15 (57).

### Brain imaging

Image acquisitions have been described in detail before (50). Put briefly, 3D T1-weighted rapid acquisition gradient-echo sequence (MPRAGE, 0.83×0.83×0.94mm), 3D T2-weighted FLAIR (0.75×0.75×0.9mm) and single-shell diffusion MRI (2×2×2mm, 64 noncollinear gradient directions, b=1000 s/mm^2^) were acquired on a single 3T Siemens Skyra MRI scanner (Siemens, Erlangen, Germany). Detailed parameters can be found in the **SI Appendix**.

An overview of the derived imaging markers for the grey and white matter can be found in **Figure 1**. For a detailed account on image preprocessing, derivation of morphometric and diffusion indices, as well as QA please refer to the **SI Appendix** (58).

Following preprocessing, we derived conventional diffusion tensor imaging (DTI) markers of white matter microstructure, i.e., FA and MD, which have been extensively used in neuroscientific and neuropsychological research (59, 60). Free-water imaging was employed to model an extracellular free-water compartment, sensitive to immune activation (61) and atrophy (62), as well as a cellular tissue compartment (FA_T_), more closely reflecting myelin and axonal alterations than their DTI equivalents (63). Fixel-based analysis, a novel multi-tissue model addressing more complex white matter compositions, was used to derive metrics of fiber density, fiber-bundle cross section (FC), fiber density and cross section (FDC), and complexity (64). For further statistical analysis, diffusion markers were averaged across a representative skeleton of the entire white matter derived by tract-based spatial statistics (65). Finally, PSMD, a surrogate marker of cerebral small vessel disease, was calculated (23).

After cortical surface reconstruction with the Computational Anatomy Toolbox for SPM (CAT12), mean cortical thickness was estimated as a proxy for neurodegenerative processes (20, 66, 67). Normalized volumes of white matter hyperintensities (WMH load) were obtained by FSL’s Brain Intensity AbNormality Classification Algorithm (BIANCA) with LOCally Adaptive Threshold Estimation (LOCATE) (68, 69).

### Statistical analysis

All statistical analyses were conducted in python 3.9.1 (70, 71), CAT12 (66, 67, 72), as well as mrclusterstats (73). Statistical tests were two-sided, with a *P*<0.05 as significance threshold. In the case of averaged imaging and clinical data, *P-*values were adjusted by Bonferroni-correction. Additionally, sensitivity analyses were performed by [1] excluding post-SARS-CoV-2 individuals who had been hospitalized, and [2] stratifying the post-SARS-CoV-2 group by recruitment strategy, following the same procedures as described below.

#### Phenotypical data

Sample characteristics were compared between healthy controls and post-SARS-CoV-2 participants using X^2^-tests (binary) and two-sample t-tests (continuous). Clinical variables were compared between groups in separate analyses of covariance (ANCOVA) adjusted for age, sex, and education.

#### Imaging

Statistical analysis of imaging parameters was conducted in two steps. First, global measures, i.e., mean skeletonized diffusion parameters, mean cortical thickness, WMH load and PSMD, were compared between post-SARS-CoV-2 individuals and healthy controls in separate ANCOVAs, adjusted for age, sex and education. In the case of FC and FDC, total intracranial volume served as an additional covariate. Next, in an effort to interrogate spatial patterns of brain structural alterations associated with a mild to moderate SARS-CoV-2 infection, we performed whole-brain voxel-wise permutation-based testing for skeletonized diffusion markers. Utilizing the same design matrices as in the ANCOVAs, we employed 5000 permutations, threshold-free cluster enhancement and family-wise error correction across multiple hypotheses. Vertex-wise cortical thickness was statistically analyzed in a general linear model as implemented in CAT12 with family-wise error correction and a cluster threshold of 10.

#### Associations between clinical and imaging data

In case of significant group differences of averaged imaging markers, we performed exploratory regression analyses testing for associations between these imaging markers and neuropsychological scores in the entire sample. Moreover, group x imaging marker interactions were included in the regression model and post-hoc Spearman correlations were conducted for each group separately. As we deemed these analyses exploratory, no correction for multiple comparisons were performed.

### Machine learning prediction

To further evaluate their predictive capacities, all brain imaging markers calculated in the study were averaged within regions of interest where applicable (Desikan-Killiany cortical atlas parcels and TractSeg-derived anatomical white matter tracts) and propagated to a comparative supervised machine learning pipeline (scikit-learn v1.0.2) (74–76). Per marker, multivariate logistic regression models were trained to predict past COVID-19. Models were scored with prediction accuracy and statistical significance was assessed via comparison to null model predictions. Further details are provided in the **SI Appendix**.

### Data availability

HCHS data is not publicly available due to privacy reasons. The analysis code is publicly available on GitHub (links can be found in the **SI Appendix)**.

## Data Availability

All data produced in the present study are available upon reasonable request to the authors.

## Acknowledgments

The authors wish to acknowledge all participants of the Hamburg City Health Study and cooperation partners, patrons and the Deanery from the University Medical Center Hamburg—Eppendorf for supporting the Hamburg City Health Study. Special thanks applies to the staff at the Epidemiological Study Center for conducting the study. The participating institutes and departments from the University Medical Center Hamburg-Eppendorf contribute all with individual and scaled budgets to the overall funding. The Hamburg City Health Study is also supported by Amgen, Astra Zeneca, Bayer, BASF, Deutsche Gesetzliche Unfallversicherung (DGUV), DIFE, the Innovative medicine initiative (IMI) under grant number No. 116074 and the Fondation Leducq under grant number 16 CVD 03., Novartis, Pfizer, Schiller, Siemens, Unilever and “Förderverein zur Förderung der HCHS e.V.”.

## Supporting information

Supporting information is available online.

## References

1. E. L. Petersen, et al., Multi-organ assessment in mainly non-hospitalized individuals after SARS-CoV-2 infection: The Hamburg City Health Study COVID programme. European Heart Journal, ehab914 (2022).

2. Y. Su, et al., Multiple early factors anticipate post-acute COVID-19 sequelae. Cell 185, 881-895.e20 (2022).

3. B. Raman, et al., Medium-term effects of SARS-CoV-2 infection on multiple vital organs, exercise capacity, cognition, quality of life and mental health, post-hospital discharge. EClinicalMedicine 31, 100683 (2021).

4. S. Mehandru, M. Merad, Pathological sequelae of long-haul COVID. Nat Immunol 23, 194–202 (2022).

5. M. Ramos-Casals, P. Brito-Zerón, X. Mariette, Systemic and organ-specific immune-related manifestations of COVID-19. Nat Rev Rheumatol 17, 315–332 (2021).

6. A. Nalbandian, et al., Post-acute COVID-19 syndrome. Nat Med 27, 601–615 (2021).

7. M. Taquet, J. R. Geddes, M. Husain, S. Luciano, P. J. Harrison, 6-month neurological and psychiatric outcomes in 236 379 survivors of COVID-19: a retrospective cohort study using electronic health records. The Lancet Psychiatry 8, 416–427 (2021).

8. A. Hampshire, et al., Cognitive deficits in people who have recovered from COVID-19. EClinicalMedicine 39, 101044 (2021).

9. J. Matschke, et al., Neuropathology of patients with COVID-19 in Germany: a post-mortem case series. The Lancet Neurology 19, 919–929 (2020).

10. O. M. Espíndola, et al., Inflammatory Cytokine Patterns Associated with Neurological Diseases in Coronavirus Disease 2019. Ann Neurol 89, 1041–1045 (2021).

11. G. Douaud, et al., SARS-CoV-2 is associated with changes in brain structure in UK Biobank. Nature, 1–17 (2022).

12. I. I. Ismail, S. Salama, Association of CNS demyelination and COVID-19 infection: an updated systematic review. J Neurol 269, 541–576 (2022).

13. M. G. Savelieff, E. L. Feldman, A. M. Stino, Neurological sequela and disruption of neuron-glia homeostasis in SARS-CoV-2 infection. Neurobiology of Disease 168, 105715 (2022).

14. J. Wenzel, et al., The SARS-CoV-2 main protease Mpro causes microvascular brain pathology by cleaving NEMO in brain endothelial cells. Nat Neurosci 24, 1522– 1533 (2021).

15. L.-A. Teuwen, V. Geldhof, A. Pasut, P. Carmeliet, COVID-19: the vasculature unleashed. Nat Rev Immunol 20, 389–391 (2020).

16. R. Manca, M. De Marco, P. G. Ince, A. Venneri, Heterogeneity in Regional Damage Detected by Neuroimaging and Neuropathological Studies in Older Adults With COVID-19: A Cognitive-Neuroscience Systematic Review to Inform the Long-Term Impact of the Virus on Neurocognitive Trajectories. Frontiers in Aging Neuroscience 13 (2021).

17. S. Kremer, et al., Neuroimaging in patients with COVID-19: a neuroradiology expert group consensus. Eur Radiol (2022) https://doi.org/10.1007/s00330-021-08499-0 (April 12, 2022).

18. S. G. Kandemirli, et al., Brain MRI Findings in Patients in the Intensive Care Unit with COVID-19 Infection. Radiology 297, E232–E235 (2020).

19. S. Katal, S. Balakrishnan, A. Gholamrezanezhad, Neuroimaging and neurologic findings in COVID-19 and other coronavirus infections: A systematic review in 116 patients. Journal of Neuroradiology 48, 43–50 (2021).

20. C. R. Jack, et al., Tracking pathophysiological processes in Alzheimer’s disease: an updated hypothetical model of dynamic biomarkers. The Lancet Neurology 12, 207–216 (2013).

21. Y. Assaf, O. Pasternak, Diffusion Tensor Imaging (DTI)-based White Matter Mapping in Brain Research: A Review. J Mol Neurosci 34, 51–61 (2008).

22. J. M. Wardlaw, C. Smith, M. Dichgans, Small vessel disease: mechanisms and clinical implications. The Lancet Neurology 18, 684–696 (2019).

23. E. Baykara, et al., A Novel Imaging Marker for Small Vessel Disease Based on Skeletonization of White Matter Tracts and Diffusion Histograms. Annals of Neurology 80, 581–592 (2016).

24. J. Meinhardt, et al., Olfactory transmucosal SARS-CoV-2 invasion as a port of central nervous system entry in individuals with COVID-19. Nat Neurosci 24, 168–175 (2021).

25. R. Butowt, K. Bilinska, SARS-CoV-2: Olfaction, Brain Infection, and the Urgent Need for Clinical Samples Allowing Earlier Virus Detection. ACS Chem. Neurosci. 11, 1200–1203 (2020).

26. M.-E. Tremblay, C. Madore, M. Bordeleau, L. Tian, A. Verkhratsky, Neuropathobiology of COVID-19: The Role for Glia. Front Cell Neurosci 14, 592214 (2020).

27. M. Schwabenland, et al., Deep spatial profiling of human COVID-19 brains reveals neuroinflammation with distinct microanatomical microglia-T-cell interactions. Immunity 54, 1594-1610.e11 (2021).

28. O. Pasternak, N. Sochen, Y. Gur, N. Intrator, Y. Assaf, Free water elimination and mapping from diffusion MRI. Magnetic Resonance in Medicine 62, 717–730 (2009).

29. O. Pasternak, et al., Excessive Extracellular Volume Reveals a Neurodegenerative Pattern in Schizophrenia Onset. J Neurosci 32, 17365–17372 (2012).

30. Y. Wang, et al., Quantification of increased cellularity during inflammatory demyelination. Brain 134, 3590–3601 (2011).

31. M. Schwartz, O. Butovsky, W. Brück, U.-K. Hanisch, Microglial phenotype: is the commitment reversible? Trends Neurosci 29, 68–74 (2006).

32. E. Syková, C. Nicholson, Diffusion in brain extracellular space. Physiol Rev 88, 1277–1340 (2008).

33. F. W. Chioh, et al., Convalescent COVID-19 patients are susceptible to endothelial dysfunction due to persistent immune activation. eLife 10, e64909 (2021).

34. S. Krasemann, et al., The Blood-Brain Barrier is Dysregulated in COVID-19 and Serves as a CNS Entry Route for SARS-CoV-2 (2021) https://doi.org/10.2139/ssrn.3828200 (June 15, 2022).

35. T. Dhollander, et al., Fixel-based Analysis of Diffusion MRI: Methods, Applications, Challenges and Opportunities. NeuroImage 241, 118417 (2021).

36. M. Petersen, et al., Fixel based analysis of white matter alterations in early stage cerebral small vessel disease. Sci Rep 12, 1581 (2022).

37. V. F. J. Newcombe, et al., Neuroanatomical substrates of generalized brain dysfunction in COVID-19. Intensive Care Med 47, 116–118 (2021).

38. Y. Qin, et al., Long-term microstructure and cerebral blood flow changes in patients recovered from COVID-19 without neurological manifestations. J Clin Invest 131, 147329 (2021).

39. S. Huang, et al., Persistent white matter changes in recovered COVID-19 patients at the 1-year follow-up. Brain, awab435 (2021).

40. L. Griffanti, et al., Adapting the UK Biobank Brain Imaging Protocol and Analysis Pipeline for the C-MORE Multi-Organ Study of COVID-19 Survivors. Frontiers in Neurology 12 (2021).

41. L. Conforti, J. Gilley, M. P. Coleman, Wallerian degeneration: an emerging axon death pathway linking injury and disease. Nat Rev Neurosci 15, 394–409 (2014).

42. A. B. Storsve, A. M. Fjell, A. Yendiki, K. B. Walhovd, Longitudinal Changes in White Matter Tract Integrity across the Adult Lifespan and Its Relation to Cortical Thinning. PLOS ONE 11, e0156770 (2016).

43. B. Besteher, et al., Larger gray matter volumes in neuropsychiatric long-COVID syndrome. Psychiatry Research 317, 114836 (2022).

44. D. R. Feikin, et al., Duration of effectiveness of vaccines against SARS-CoV-2 infection and COVID-19 disease: results of a systematic review and meta-regression. The Lancet 399, 924–944 (2022).

45. A. Carfì, R. Bernabei, F. Landi, for the Gemelli Against COVID-19 Post-Acute Care Study Group, Persistent Symptoms in Patients After Acute COVID-19. JAMA 324, 603–605 (2020).

46. C. Huang, et al., 6-month consequences of COVID-19 in patients discharged from hospital: a cohort study. The Lancet 397, 220–232 (2021).

47. Z. Al-Aly, Y. Xie, B. Bowe, High-dimensional characterization of post-acute sequelae of COVID-19. Nature 594, 259–264 (2021).

48. A. Jagodzinski, U. Koch-gromus, G. Adam, S. Anders, M. Augustin, Rationale and Design of the Hamburg City Health Study. European Journal of Epidemiology (2019) https://doi.org/10.1007/s10654-019-00577-4.

49. D. Ho, K. Imai, G. King, E. A. Stuart, MatchIt: Nonparametric Preprocessing for Parametric Causal Inference. Journal of Statistical Software 42, 1–28 (2011).

50. M. Petersen, et al., Network Localisation of White Matter Damage in Cerebral Small Vessel Disease. Scientific Reports 10, 9210 (2020).

51. M. F. Folstein, S. E. Folstein, P. R. McHugh, “Mini-mental state”. A practical method for grading the cognitive state of patients for the clinician. J Psychiatr Res 12, 189– 198 (1975).

52. T. Tombaugh, Trail Making Test A and B: Normative data stratified by age and education. Archives of Clinical Neuropsychology 19, 203–214 (2004).

53. J. C. Moms, et al., The Consortium to Establish a Registry for Alzheimer’s Disease (CERAD). Part I. Clinical and neuropsychological assesment of Alzheimer’s disease. Neurology 39, 1159–1159 (1989).

54. K. I. Shulman, Clock-drawing: is it the ideal cognitive screening test? International Journal of Geriatric Psychiatry 15, 548–561 (2000).

55. R. L. Spitzer, K. Kroenke, J. B. W. Williams, B. Löwe, A Brief Measure for Assessing Generalized Anxiety Disorder: The GAD-7. Arch Intern Med 166, 1092 (2006).

56. K. Kroenke, R. L. Spitzer, J. B. W. Williams, The PHQ-9: Validity of a brief depression severity measure. J Gen Intern Med 16, 606–613 (2001).

57. K. Kroenke, R. L. Spitzer, J. B. W. Williams, The PHQ-15: Validity of a New Measure for Evaluating the Severity of Somatic Symptoms: Psychosomatic Medicine 64, 258– 266 (2002).

58. M. Cieslak, et al., QSIPrep: an integrative platform for preprocessing and reconstructing diffusion MRI data. Nat Methods 18, 775–778 (2021).

59. R. J. Cannistraro, et al., CNS small vessel disease: A clinical review. Neurology 92, 1146–1156 (2019).

60. K. Kamagata, et al., Diffusion Magnetic Resonance Imaging-Based Biomarkers for Neurodegenerative Diseases. Int J Mol Sci 22, 5216 (2021).

61. M. A. Di Biase, et al., Large-Scale Evidence for an Association Between Peripheral Inflammation and White Matter Free Water in Schizophrenia and Healthy Individuals. Schizophr Bull 47, 542–551 (2021).

62. E. Ofori, et al., Free-water imaging of the hippocampus is a sensitive marker of Alzheimer’s disease. NeuroImage: Clinical 24 (2019).

63. O. Pasternak, N. Sochen, Y. Gur, N. Intrator, Y. Assaf, Free water elimination and mapping from diffusion MRI. Magnetic Resonance in Medicine 62, 717–730 (2009).

64. T. Dhollander, et al., Fixel-based Analysis of Diffusion MRI: Methods, Applications, Challenges and Opportunities. NeuroImage 241, 118417 (2021).

65. S. M. Smith, et al., Tract-based spatial statistics: Voxelwise analysis of multi-subject diffusion data. NeuroImage 31, 1487–1505 (2006).

66. R. Dahnke, R. A. Yotter, C. Gaser, Cortical thickness and central surface estimation. NeuroImage 65, 336–348 (2013).

67. C. Gaser, et al., CAT – A Computational Anatomy Toolbox for the Analysis of Structural MRI Data. 2022.06.11.495736 (2022).

68. L. Griffanti, et al., BIANCA (Brain Intensity AbNormality Classification Algorithm): A new tool for automated segmentation of white matter hyperintensities. NeuroImage 141, 191–205 (2016).

69. V. Sundaresan, et al., Automated lesion segmentation with BIANCA: Impact of population-level features, classification algorithm and locally adaptive thresholding. NeuroImage 202, 116056 (2019).

70. R. Vallat, Pingouin: statistics in Python. JOSS 3, 1026 (2018).

71. P. Virtanen, et al., SciPy 1.0: fundamental algorithms for scientific computing in Python. Nat Methods 17, 261–272 (2020).

72. R. A. Yotter, R. Dahnke, P. M. Thompson, C. Gaser, Topological correction of brain surface meshes using spherical harmonics. Hum. Brain Mapp. 32, 1109–1124 (2011).

73. J.-D. Tournier, et al., MRtrix3: A fast, flexible and open software framework for medical image processing and visualisation. NeuroImage 202, 116137 (2019).

74. A. Abraham, et al., Machine learning for neuroimaging with scikit-learn. Frontiers in Neuroinformatics 8 (2014).

75. R. S. Desikan, et al., An automated labeling system for subdividing the human cerebral cortex on MRI scans into gyral based regions of interest (2006).

76. J. Wasserthal, P. Neher, K. H. Maier-Hein, TractSeg - Fast and accurate white matter tract segmentation. NeuroImage 183, 239–253 (2018).

