## Supplementary materials for "Brain imaging and neuropsychological assessment of individuals recovered from a mild to moderate SARS-CoV-2 infection"

–

*Supplemental Material*

Marvin Petersen, MD[1#]; Felix Leonard Nägele, MD[1#]; Carola Mayer, MSc[1]; Maximilian Schell[1]; Elina Petersen, MSc[2,3]; Simone Kühn, PhD[4]; Jürgen Gallinat, MD[4]; Jens Fiehler, MD[5]; Ofer Pasternak, PhD[6]; Jakob Matschke, MD[7]; Markus Glatzel, MD[7]; Raphael Twerenbold, MD[2,3,8,9]; Christian Gerloff, MD[1]; Götz Thomalla, MD[1]; Bastian Cheng, MD[1]

[1]Department of Neurology, University Medical Center Hamburg-Eppendorf, Hamburg, Germany

[2]Department of Cardiology, University Heart and Vascular Center, Hamburg, Germany

[3]Population Health Research Department, University Heart and Vascular Center, Hamburg, Germany

[4]Department of Psychiatry and Psychotherapy, University Medical Center Hamburg-Eppendorf, Hamburg, Germany

[5]Department of Neuroradiology, University Medical Center Hamburg-Eppendorf, Hamburg, Germany

[6]Department of Psychiatry and Radiology, Brigham and Women's Hospital, Harvard Medical School, Boston, MA, USA

[7]Institute of Neuropathology, University Center Hamburg-Eppendorf, Hamburg, Gemany

[8]German Center for Cardiovascular Research (DZHK), partner site Hamburg/Kiel/Luebeck, Hamburg, Germany

[9]University Center of Cardiovascular Science, University Heart and Vascular Center, Hamburg, Germany

**# these authors contributed equally**

### Methods

#### Image acquisition

Image acquisitions were conducted on a single 3T Siemens Skyra MRI scanner (Siemens, Erlangen, Germany). 3D T1-weighted rapid acquisition gradient-echo sequence (MPRAGE): repetition time (TR) = 2500 ms, echo time (TE) = 2.12 ms, 256 axial slices, slice thickness (ST) = 0.94 mm, and in-plane resolution (IPR) = 0.83 x 0.83 mm; 3D T2-weighted FLAIR: TR = 4700 ms, TE = 392 ms, 192 axial slices, ST = 0.9 mm, and IPR = 0.75 x 0.75 mm; and single-shell diffusion MRI: TR = 8500 ms, TE = 75 ms, 75 axial slices, ST = 2 mm, IPR = 2 x 2mm, 64 noncollinear gradient directions with b = 1000 s/mm$^2$, 1 image with b = 0 s/mm$^2$.

#### Image processing

##### T1-weighted MRI

###### *Preprocessing*

Preprocessing was performed using *QSIPrep* 0.14.2[1], which is based on *Nipype* 1.6.1[2]. The T1-weighted (T1w) image was corrected for intensity non-uniformity (INU) using N4BiasField-Correction[3] (ANTs 2.3.1), and used as T1w-reference throughout the workflow. The T1w-reference was then skull-stripped using antsBrainExtraction.sh (ANTs 2.3.1), using OASIS as target template.

###### *Estimation of cortical thickness*

Surface-based morphometry was conducted in the *Computational Anatomy Toolbox* for *SPM* (*CAT12*)[5] for cortical surface reconstruction and estimation of mean cortical thickness employing the projection-based thickness method[6], as well as topology correction[7] and spherical mapping[8].

##### Diffusion-weighted MRI

###### *Preprocessing*

*QSIPrep* 0.14.2[1] was also used for preprocessing of diffusion-weighted MRI (dMRI). MP-PCA denoising as implemented in MRtrix3's dwidenoise[9] was applied with a 5-voxel window. After

MP-PCA, Gibbs unringing was performed using MRtrix3's mrdegibbs.[10] Following unringing, B1 field inhomogeneity was corrected using dwibiascorrect from MRtrix3 with the N4 algorithm.[3]

FSL (version 6.0.3:b862cdd5)'s eddy was used for head motion correction and eddy current correction.[11] Eddy was configured with a $q$-space smoothing factor of 10, a total of 5 iterations, and 1000 voxels used to estimate hyperparameters. A linear first level model and a linear second level model were used to characterize eddy current-related spatial distortion. $q$-space coordinates were forcefully assigned to shells. Field offset was attempted to be separated from subject movement. Shells were aligned post-eddy. Eddy's outlier replacement was run. Data were grouped by slice, only including values from slices determined to contain at least 250 intracerebral voxels. Groups deviating by more than 4 standard deviations from the prediction had their data replaced with imputed values. Final interpolation was performed using the jac method.

A deformation field to correct for susceptibility distortions was estimated based on fMRIprep's fieldmap-less approach.[12] The deformation field is results from co-registering the b0 reference to the same-subject T1w-reference with its intensity inverted[13]. Registration was performed with antsRegistration (ANTs 2.3.1), and the process regularized by constraining deformation to be nonzero only along the phase-encoding direction and modulated with an average fieldmap template. Based on the estimated susceptibility distortion, an unwarped b=0 reference was calculated for a more accurate co-registration with the anatomical reference. Several confounding time-series were calculated based on the preprocessed DWI: framewise displacement using the implementation in *Nipype* (following the definitions by [14]). The head-motion estimates calculated in the correction step were also placed within the corresponding confounds file. Slicewise cross correlation was also calculated. The DWI time-series were resampled to ACPC, generating a preprocessed DWI run in ACPC space with 2mm isotropic voxels.

Many internal operations of *QSIPrep* use *Nilearn*[15] and *Dipy*[16]. For more details of the pipeline, see the section corresponding to workflows in *QSIPrep*'s documentation.

#### *Diffusion tensor imaging and free-water imaging*

Fractional anisotropy (FA) and mean diffusivity (MD) were derived from diffusion tensors which were modelled based on preprocessed dMRI using a least-squares fit.[17,18] Further, we employed free-water imaging, a two tensor model, modelling an extracellular compartment of isotropic diffusion, as well as a cellular compartment characterized by hindered/restricted diffusion.[19] Thus, by means of a regularized non-linear fit, free-water, and free-water corrected diffusion tensors were estimated for each study participant from which FA of the tissue compartment was calculated ($FA_T$).[19]

#### *Fixel-based analysis pipeline*

*MRtrix3* (v.3.0.2)[20] was utilized to estimate fiber density (FD), fiber cross section (FC), fiber density and cross section (FDC), and complexity (CX) at the voxel-level.

First, the preprocessed DWI was upsampled to a voxel size of 1.25 x 1.25 x 1.25 $mm^3$, after which multi-tissue fiber response functions were estimated using the dhollander algorithm.[21] Fiber orientation distributions (FODs) were subsequently estimated via constrained spherical deconvolution[22] (CSD) using an unsupervised single-shell-optimized multi-tissue method (*MRtrix3Tissue* (https://3Tissue.github.io).[23,24] FODs were intensity-normalized using mtnormalize[25] after which a study-specific unbiased FOD template based on 20 healthy controls and 20 post-SARS-CoV-2 individuals was created with *MRtrix3*'s population_template function. Next, individual FOD images and brain masks were non-linearly registered to the white matter FOD template. Transformed brain masks were used to compute a template mask, i.e. the intersection of all subject masks in template space. In the next step, fixels (= fiber populations within a voxel) were segmented from the FOD template within the template mask, resulting in a template fixel mask which was further refined to respect crossing fibers while excluding false positives (empirically derived crossing fiber and false positive thresholds: 0.06 and 0.18, respectively).

Next, following probabilistic whole-brain tractography based on the FOD template[26] (angle 22.5, maxlen 250, minlen 10, power 1, 20 x $10^6$ streamlines, cutoff 0.06) and spherical-deconvolution informed filtering of whole-brain tractograms[27], deep learning based tract segmentation was performed with TractSeg[28]. The resulting tract segmentations were utilized to extract averaged diffusion indices for 72 major white matter tracts which served as features for logistic regression models predicting group membership.

Moreover, the transformed individual FOD images were segmented to derive fixels and their apparent FD.[29] Then, fixels of all subjects in template space were reoriented based on the local transformation at each voxel in the warps used previously. Subsequently, each subject's fixels were assigned to template fixels enabling statistical analysis of common, i.e., corresponding, fiber populations. FC was derived from non-linear warps generated during registration of individual FODs to template space after which the logarithm of FC (Log. FC) was calculated to ensure a zero centered normal distribution.[29] FDC was calculated as the product of FC and FD. Based on the whole-brain streamlines tractogram, a fixel-fixel connectivity matrix was computed which was then used for smoothing the fixel metrics FD, Log. FC and FDC. In order to derive fixel metrics on the voxel-level, they were averaged across all fixels within a voxel using MRtrix3's fixel2voxel function. Moreover, CX, a metric of crossing-fiber organization, was calculated.[30]

In an effort to allow for comparisons with conventional diffusion tensor imaging and free-water imaging markers, a study specific FA template was created. Therefore, previously derived individual non-linear warps from native FOD to FOD template space were used to register FA maps to FOD template space. These FA maps in FOD template space were then averaged and the resulting study-specific FA template served as the registration target for non-linear transformations of FA images from native space to template space utilizing ANTs' SyN registration.[31] The resulting transformations were subsequently applied to the remaining maps of diffusion tensor imaging and free-water imaging metrics.

##### *Tract-based spatial statistics*

In order to derive skeletonized maps of each of the estimated diffusion parameters, we conducted tract-based spatial statistics (*TBSS*)[32,33] utilizing the above described study-specific FA template as the registration target. Briefly, individual FA images in template space got eroded to exclude non-brain voxels on the outer edge of the image. Next, a valid mask containing only the intersection of all subjects' brains was derived and used to mask the average of all previously eroded FA images. This mean FA image was subsequently used to derive a white matter skeleton which was thresholded at FA > 0.25. Next, all individual FA images were projected onto the mean FA skeleton. The resultant projection vectors were used to skeletonize all of the remaining diffusion metrics including those from free-water imaging and fixel-based analysis pipelines. Finally, diffusion markers were averaged across the entire white matter skeleton for further statistical analysis.

##### *Peak width of skeletonized mean diffusivity*

Peak width of skeletonized mean diffusivity (PSMD) was calculated based on standard procedures[34] adapted in terms of the non-linear registration step for which we used *ANTs*' SyN registration.[31] PSMD is calculated as the difference between the 95$^{th}$ and 5$^{th}$ percentile of MD values on the white matter skeleton in standard (MNI) space. A mask supplied by the developers was used to exclude white matter areas susceptible to partial volume effects of cerebrospinal fluid (https://github.com/miac-research/psmd/blob/main/skeleton_mask_2019.nii.gz).

#### Fluid-attenuated inversion recovery MRI

##### *White matter hyperintensity segmentation*

*FSL*'s *Brain Intensity AbNormality Classification Algorithm* (*BIANCA*)[35] with *LOCally Adaptive Threshold Estimation* (*LOCATE*)[36] were applied on FLAIR images and T1w images for white matter hyperintensity (WMH) segmentation.

The training dataset for the supervised k-nearest neighbor algorithm (*BIANCA*) comprised nearly 100 WMH masks manually segmented on FLAIR images by two independent raters. The manual segmentations of both raters were inclusively added into one binary mask for each participant and served as the training dataset for *BIANCA*[35] and *LOCATE*[36]. For the training of *BIANCA*[35], the following images were used as input: 1) a brain-extracted FLAIR image; 2) a transformation matrix based on a linear registration from FLAIR space to standard MNI space (MNI152NLin2009cAsym template) utilizing *FSL's FLIRT* tool;[37,38] 3) a T1w image rigidly registered to FLAIR space with *AntsRegistration;*[39] 4) the manually segmented WMH masks. We used 3D patches, selected the non-lesion points from "no border", and chose 2.000 training points and 10.000 non-lesion points as segmentation parameters. Based on the initial training of *BIANCA*[35], the testing dataset was automatically segmented using the same input images and parameters but without manual segmentations. Participants included in the training dataset were segmented in a leave-one-out validation by defining the query subject parameter in *BIANCA.*[35]

The raw output masks of *BIANCA*[35] were used as input for *LOCATE*[36]. In addition to the *BIANCA*[35] masks, *LOCATE*[36] further received as input brain-extracted FLAIR and rigidly registered T1-weighted images in FLAIR space, as also used in *BIANCA*[35]*.* Moreover, further inputs were a ventricle distancemap created based on *Freesurfer v.7.1* output*,*[40] the manual segmentations for training and a brain mask in FLAIR space. After training with *LOCATE*[36], participants included in the training dataset were again segmented with a leave-one-out validation. The remaining participants in the testing dataset were automatically segmented.

After application of the *BIANCA*[35] and *LOCATE*[36] algorithms*,* the segmentations were further refined using *Freesurfer v.7.1* parcellations[40] to exclude non-white matter regions. Specifically, a dilated cortical ribbon mask, an eroded ventricle mask, and parcellations from the corpus callosum and basal ganglia were used as exclusion masks. Lesion clusters were filtered for a minimum cluster size of 5 voxels as defined by the 6-connectivity. Finally, the lesion load was retrieved after normalizing for intracranial volume, as calculated by *Freesurfer v.7.1.*[40]

#### Quality assurance

Quality assurance (QA) of MRI data was conducted both quantitatively and qualitatively. First, a neuroradiologist reviewed all imaging data for pathologies. Further, for raw data, quantitative

QA measures were derived for T1w and dMRI data utilizing *MRIQC*[41], *Freesurfer*[42] and *QSI-Prep.*[1] Qualitative QA of raw imaging data was subsequently performed for outliers in frame-wise displacement and number of slices with signal dropouts (dMRI), as well as cortical thickness, brain volumes and coefficient of joint variation (T1w), in each case defined by ± 2 standard deviations from the mean. The quality of FLAIR images was assessed visually. Last, all derivatives of neuroimaging pipelines were visually assessed in order to ensure appropriate processing.

#### Machine learning prediction

To further evaluate the predictive capacity of derived imaging markers, they separately served as input to a comparative supervised machine learning pipeline. Therefore, average cortical thickness within Desikan-Killiany atlas parcels[43] was computed and diffusion markers were averaged within predefined anatomical fiber tracts from TractSeg outputs.[28] Per marker, multivariate logistic regression models were trained to predict whether a participant has COVID-19. Elastic net penalties[44] were applied for model regularization and SAGA served as the underlying optimization algorithm[45]. As a binary categorization task was performed, models were scored using accuracy which is also the metric we report.

$$Accuracy = \frac{TP + TN}{TP + TN + FP + FN}$$

Where TP = true positive, TN = true negative, FP = false positive, FN = false negative.

Accuracy scores provide an intuitive measure facilitating between-marker comparison and evaluation of a marker's diagnostic merit beyond abstract effect sizes derived from inferential statistics. Model training, corresponding parameter optimization and evaluation were conducted in a 10-fold nested cross-validation setup (L1-ratios=0.1,0.5,0.7,0.9,0.95,0.99,1, $n_{Cs}$=10) to prevent data leakage and consequent overfitting. Optimization procedures were repeated 100 times with different random split regimens in the cross-validation to make sure that prediction results were not biased by a single arbitrary split.[46] To assess whether prediction performance was statistically significant median accuracy scores obtained from aforementioned analysis were compared to the accuracy distribution of null prediction models where group membership was randomly permuted ($n_{permutations}$ = 1000). The prediction analysis was performed using scikit-learn (v1.0.2).[15]

### Results

#### Main analyses

**Figure S1. Matching results visualized as a balance plot**

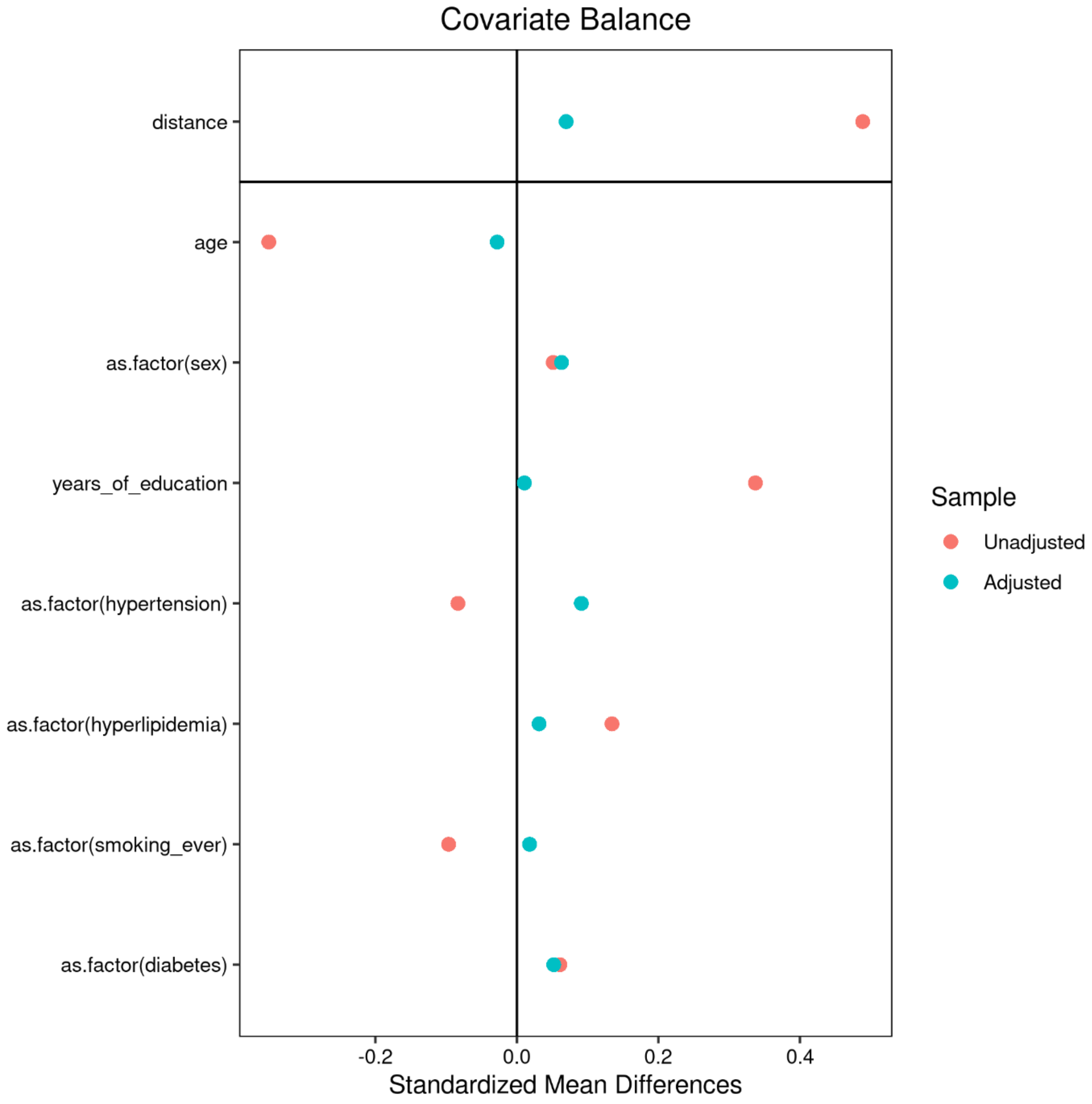

Figure S1 shows the standardized mean differences between the healthy control and post-SARS-CoV-2 groups for each matching variable before (unadjusted, red) and after matching (adjusted, turquoise). The closer the standardized mean difference is to zero, the more similar the groups are. Each matching variable is depicted separately, i.e., from top to bottom, age, sex, years of education, hypertension, hyperlipidemia, smoking and diabetes.

**Table S1. Results of analyses of covariance comparing averaged imaging markers between post-SARS-CoV-2 individuals and matched controls**

| Imaging metric[a] | Post-SARS-CoV-2 | Matched controls | $P_{uncorr}$[b] | $P_{bonf}$[c] | *F* |
|---|---|---|---|---|---|
| **FA** | 0.480 ± 0.016 (221) | 0.482 ± 0.016 (206) | .20 | >.99 | 1.63 |
| **MD ($10^{-3}$ $mm^2$/s)** | 0.747 ± 0.021 (221) | 0.740 ± 0.020 (206) | <.001 | **<.001*** ** | 17.28 |
| **$FA_T$** | 0.566 ± 0.010 (221) | 0.564 ± 0.011 (206) | .06 | .61 | 3.69 |
| **FW** | 0.148 ± 0.018 (221) | 0.142 ± 0.017 (206) | <.001 | **<.001*** ** | 18.47 |
| **FD** | 0.526 ± 0.052 (219) | 0.531 ± 0.031 (203) | .18 | >.99 | 1.84 |
| **FDC** | 0.540 ± 0.076 (219) | 0.551 ± 0.055 (203) | .11 | >.99 | 2.62 |
| **Log. FC** | 0.008 ± 0.189 (219) | 0.016 ± 0.198 (205) | .80 | >.99 | .06 |
| **CX** | 0.633 ± 0.029 (219) | 0.634 ± 0.020 (203) | .63 | >.99 | .23 |
| **PSMD ($10^{-3}$ $mm^2$/s)** | 0.212 ± 0.031 (221) | 0.207 ± 0.023 (206) | .005 | .053 | 8.03 |
| **WMH Load (%)** | 0.105 ± 0.150 (205) | 0.099 ± 0.122 (207) | .39 | >.99 | .73 |
| **CT (mm)** | 2.571 ± 0.097 (221) | 2.550 ± 0.094 (221) | .01 | .12 | 6.52 |

*Abbreviations*: CT = cortical thickness, CX = complexity, FA = fractional anisotropy, $FA_T$ = FA of the tissue, FD = fiber density, FDC = fiber density and cross-section, FW = free-water, Log. FC = logarithm of fiber cross-section, MD = mean diffusivity, post-SARS-CoV-2 = individuals who recovered from a severe acute respiratory coronavirus type 2 infection, PSMD = peak width of skeletonized MD, WMH = white matter hyperintensity

[a]Presented as mean ± standard deviation (N)
[b]Uncorrected *P* values of analyses of covariance, adjusted for age, sex and years of education
[c]Bonferroni-corrected *P* values of analyses of covariance, adjusted for age, sex and years of education (considering 11 comparisons)
***Denotes statistical significance at Bonferroni-corrected *P* <.001

**Table S2. Results of white matter voxel-wise statistics comparing post-SARS-CoV-2 individuals with matched controls**

| Imaging metric | ***All*** **post-SARS-CoV-2 (N = 221)** | ***Non-hospitalized*** **post-SARS-CoV-2 (N = 203)** |
|---|---|---|
| | **Percentage of significant voxels ($P_{FWE}$ < .05)** | |
| ***Post-SARS-CoV-2 individuals > matched controls*** | | |
| **FA** | 0.8 | 0.6 |
| **MD** | 41.3 | 40.5 |
| **$FA_T$** | 3.3 | 2.9 |
| **FW** | 38.3 | 38.0 |
| **FD** | 0 | 0 |
| **FDC** | <.01 | <.01 |
| **Log. FC** | 0 | 0 |
| **CX** | 0 | 0 |
| ***Post-SARS-CoV-2 < matched controls*** | | |
| **FA** | 1.2 | 1.4 |
| **MD** | 1.0 | 0.9 |
| **$FA_T$** | 0 | 0 |
| **FW** | 0.4 | 0.4 |
| **FD** | 0 | 0 |
| **FDC** | 2.5 | 2.7 |
| **Log. FC** | 0.7 | 0.4 |
| **CX** | <.1 | <.1 |

*Abbreviations*: CX = complexity, FA = fractional anisotropy, $FA_T$ = FA of the tissue, FD = fiber density, FDC = fiber density and cross-section, FW = free-water, FWE = family-wise error corrected, Log. FC = logarithm of fiber cross-section, MD = mean diffusivity, post-SARS-CoV-2 = individuals who recovered from a severe acute respiratory coronavirus type 2 infection

#### Exploratory analyses

##### Correlations of clinical measures with free-water

**Table S3. Results of exploratory regression analyses and post-hoc spearman correlations, associating average free-water with clinical outcome measures.**

| Outcome | Intercept (SE), *P* | FW (SE), *P* | FW:Group (SE), *P* | Post-SARS-CoV-2 Spearman *rho*, *P* | Matched Controls Spearman *rho*, *P* |
|---|---|---|---|---|---|
| **Neurocognition** | | | | | |
| **TMT-A** | 21.39 (4.54), **<.001*** ** | 85.87 (32.10), **.008** ** | -15.29 (7.79), .05 | 0.20, **.004** ** | 0.14, .07 |
| **TMT-B** | 32.92 (9.87), **.001*** ** | 266.36 (69.64), **<.001*** ** | -270.05 (16.85), .11 | 0.22, **.001*** ** | 0.15, **.04*** |
| **VF** | 34.64 (2.69), **<.001*** ** | -57.36 (19.04), **.003** ** | 12.51 (4.57), **.006** ** | -0.23, **<.001*** ** | -0.06, .43 |
| **WLR** | 11.28 (0.67), **<.001*** ** | -21.26 (4.73), **<.001*** ** | 2.60 (1.13), **.02*** | -0.25, **<.001*** ** | -0.12, .10 |
| **MMSE** | 28.71 (0.60), **<.001*** ** | -45.49 (4.22), .28 | 2.39 (1.01), **.02*** | <.001, >.99 | -0.06, .43 |
| **CDT** | 6.44 (0.38), **<.001*** ** | 0.73 (2.66), 0.79 | 1.35 (0.64), **.04*** | 0.11, .10 | <.001, >.99 |
| **Psychosocial symptom burden** | | | | | |
| **PHQ-9** | 5.64 (1.56), **<.001*** ** | -128.35 (11.01), .25 | 14.72 (2.61), .57 | 0.03, .65 | -0.04, .57 |
| **GAD-7** | 3.40 (1.32), .01* | -45.80 (9.37), .63 | 15.64 (2.22), .48 | -.003, .97 | -.004, .95 |
| **Neurological symptom burden** | | | | | |
| **PHQ-15[a]** | 2.26 (0.73), **.003** ** | -30.81 (5.19), .55 | 2.21 (1.23), .07 | 0.02, .69 | -0.01, .85 |

*Abbreviations*: CDT = clock drawing test, FW = free-water, GAD = General Anxiety Disorder, MMSE = Mini Mental State Examination, PHQ = Patient Health Questionnaire, post-SARS-CoV-2 individuals = individuals who recovered from a severe acute respiratory coronavirus type 2 infection, SE = standard error, TMT-A = Trail-Making-Test Part A, TMT-B = TMT Part B, VF = verbal fluency, WLR = word list recall

**P* <.05, ***P* <.01, ****P* <.001, uncorrected for multiple comparisons; [a]PHQ-15 items: headache, dizziness, fatigue, sleep disturbances

**Figure S2. Scatterplots and linear regressions between clinical measures and free-water**

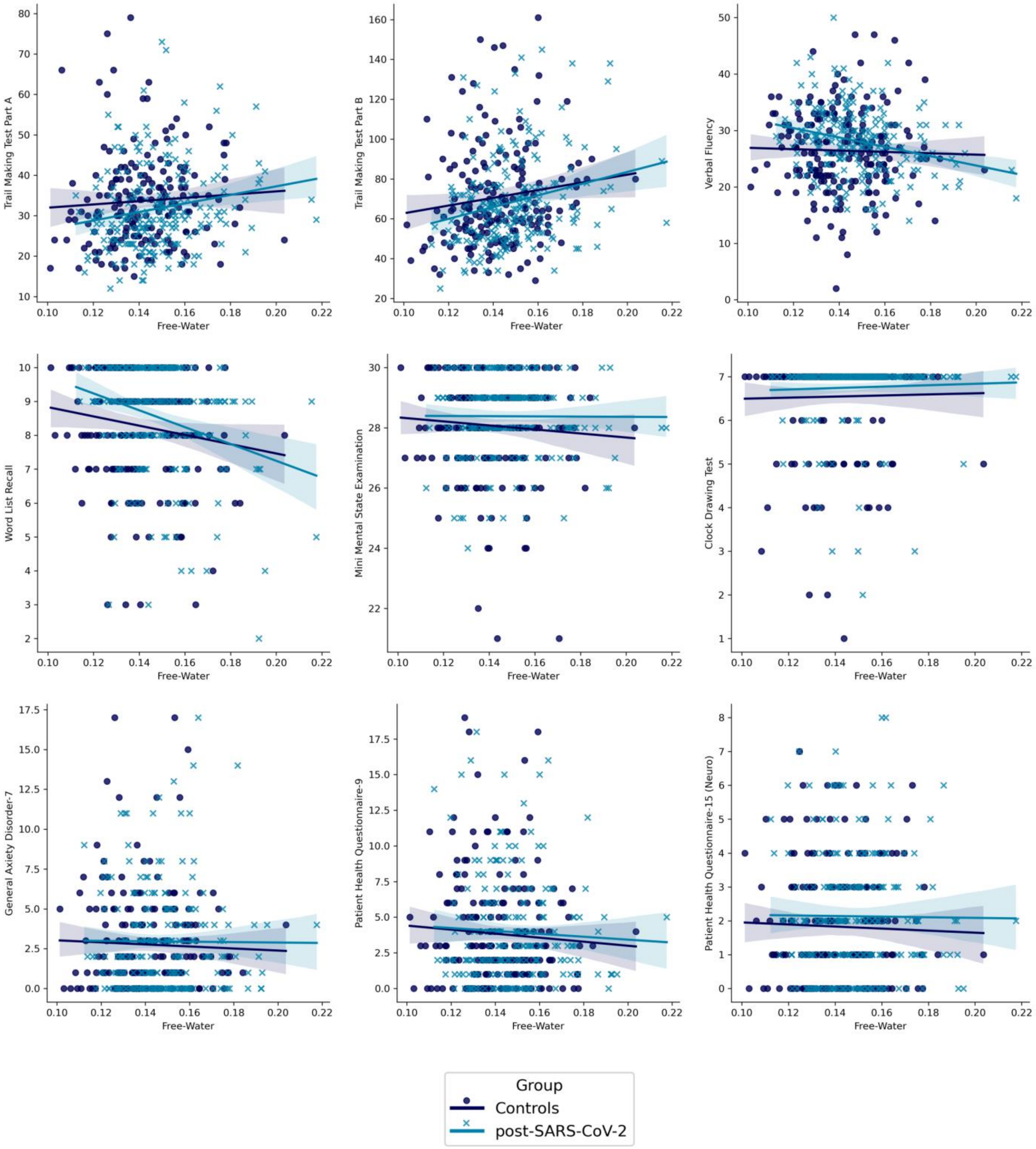

**Figure S2** shows individual data points (clinical measure vs. free-water) of matched controls and post-SARS-CoV-2 individuals in scatterplots along with a linear fit performed for each group separately. Quantitative results are displayed in **Table S3.**

#### Correlations of clinical measures with mean diffusivity

**Table S4. Results of exploratory regression analyses and post-hoc spearman correlations, associating average mean diffusivity with clinical outcome measures.**

| **Outcome** | **Intercept (SE), *P*** | **MD (SE), *P*** | **MD:Group (SE), *P*** | **Post-SARS-CoV-2 Spearman *rho*, *P*** | **Matched Controls Spearman *rho*, *P*** |
|---|---|---|---|---|---|
| **Neurocognition** | | | | | |
| **TMT-A** | -10.46 (19.66), .60 | 59615.87 (26581.83), **.03*** | -3086.31 (1528.34), **.04*** | 0.17, **.01*** | 0.13, .09 |
| **TMT-B** | -76.01 (42.73), .08 | 198475.07 (57761.53), **.001**** | -5373.59 (3311.54), .11 | 0.20, **.005**** | 0.14, .06 |
| **VF** | 55.96 (11.67), **<.001***** | -39915.30 (15774.55), **.01*** | 2572.61 (895.77), **.004**** | -0.22, **.001**** | -0.04, .60 |
| **WLR** | 20.08 (2.89), **<.001***** | -15977.89 (3911.41), **<.001***** | 520.06 (222.00), **.02*** | -0.23, **<.001***** | -0.10, .16 |
| **MMSE** | 30.21 (2.58), **<.001***** | -2899.39 (3495.06), .41 | 456.24 (198.55), **.02*** | -0.003, .97 | -0.05, .47 |
| **CDT** | 5.58 (1.63), **.001**** | 1310.07 (2201.92), .60 | 263.64 (125.71), **.04*** | 0.13, .07 | <.001, >.99 |
| **Psychosocial symptom burden** | | | | | |
| **PHQ-9** | 13.11 (6.76), .05 | -12555.57 (9141.94), .17 | 303.44 (510.00), .55 | 0.02, .76 | -0.05, .52 |
| **GAD-7** | 6.25 (5.75), .28 | -4730.24 (7785.42), .54 | 311.72 (434.33), .47 | -0.005, .94 | -0.009, .90 |
| **Neurological symptom burden** | | | | | |
| **PHQ-15[a]** | 3.36 (3.19), .29 | -2071.37 (4314.39), .63 | 432.40 (241.42), .07 | 0.02, .76 | -0.01, .89 |

*Abbreviations*: CDT = clock drawing test, MD = mean diffusivity, GAD = General Anxiety Disorder, MMSE = Mini Mental State Examination, PHQ = Patient Health Questionnaire, post-SARS-CoV-2 individuals = individuals who recovered from a severe acute respiratory coronavirus type 2 infection, SE = standard error, TMT-A = Trail-Making-Test Part A, TMT-B = TMT Part B, VF = verbal fluency, WLR = word list recall

**P* <.05, ***P* <.01, ****P* <.001, uncorrected for multiple comparisons; [a]PHQ-15 items: headache, dizziness, fatigue, sleep disturbances

**Figure S3. Scatterplots and linear regressions between clinical measures and mean-diffusivity**

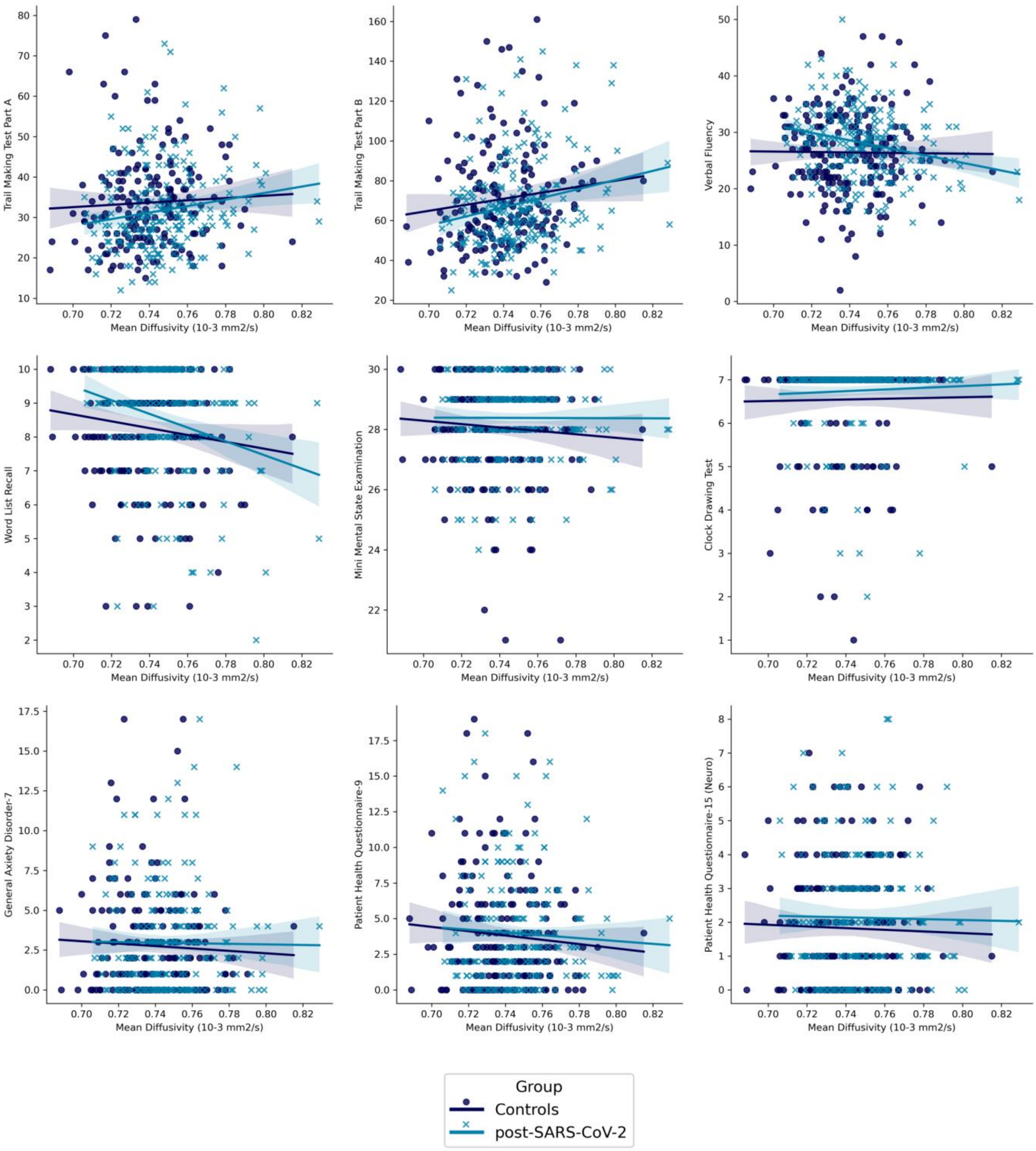

**Figure S3** shows individual data points (clinical measure vs. free-water) of matched controls and post-SARS-CoV-2 individuals in scatterplots along with a linear fit performed for each group separately. Quantitative results are displayed in **Table S4.**

### Sensitivity analyses

#### Comparison of matched controls with non-hospitalized post-SARS-CoV-2 individuals

**Table S5. Results of analyses of covariance comparing averaged imaging markers between non-hospitalized post-SARS-CoV-2 individuals and matched controls**

| Imaging metric[a] | Non-hospitalized Post-SARS-CoV-2 | Matched controls | $P_{uncorr}$[b] | $P_{bonf}$[c] | $F$ |
|---|---|---|---|---|---|
| **FA** | 0.480 ± 0.016 (203) | 0.482 ± 0.016 (206) | .17 | >.99 | 1.88 |
| **MD ($10^{-3}$ $mm^2/s$)** | 0.747 ± 0.021 (203) | 0.740 ± 0.020 (206) | <.001 | **<.001*** ** | 16.79 |
| **$FA_T$** | 0.566 ± 0.010 (203) | 0.564 ± 0.011 (206) | .09 | >.99 | 2.85 |
| **FW** | 0.148 ± 0.018 (203) | 0.142 ± 0.017 (206) | <.001 | **<.001*** ** | 17.82 |
| **FD** | 0.525 ± 0.053 (202) | 0.531 ± 0.031 (203) | .14 | >.99 | 2.14 |
| **FDC** | 0.538 ± 0.077 (202) | 0.551 ± 0.055 (203) | .09 | >.99 | 2.87 |
| **Log. FC** | 0.004 ± 0.196 (202) | 0.016 ± 0.198 (205) | .74 | >.99 | .11 |
| **CX** | 0.634 ± 0.030 (202) | 0.634 ± 0.020 (203) | .74 | >.99 | .11 |
| **PSMD ($10^{-3}$ $mm^2/s$)** | 0.210 ± 0.031 (203) | 0.207 ± 0.023 (206) | .01 | .13 | 6.43 |
| **WMH Load (%)** | 0.107 ± 0.155 (187) | 0.099 ± 0.122 (207) | .25 | >.99 | 1.32 |
| **CT (mm)** | 2.571 ± 0.097 (203) | 2.550 ± 0.094 (221) | .03 | .37 | 4.54 |

*Abbreviations*: CT = cortical thickness, CX = complexity, FA = fractional anisotropy, $FA_T$ = FA of the tissue, FD = fiber density, FDC = fiber density and cross-section, FW = free-water, Log. FC = logarithm of fiber cross-section, MD = mean diffusivity, post-SARS-CoV-2 = individuals who recovered from a severe acute respiratory coronavirus type 2 infection, PSMD = peak width of skeletonized MD, WMH = white matter hyperintensity

[a]Presented as mean ± standard deviation (N)
[b]Uncorrected *P* values of analyses of covariance, adjusted for age, sex and years of education
[c]Bonferroni-corrected *P* values of analyses of covariance, adjusted for age, sex and years of education (considering 11 comparisons)
***Denotes statistical significance at bonferroni-corrected *P* <.001

**Figure S4. Boxplots and statistics of averaged imaging markers comparing non-hospitalized post-SARS-CoV-2 individuals with matched controls**

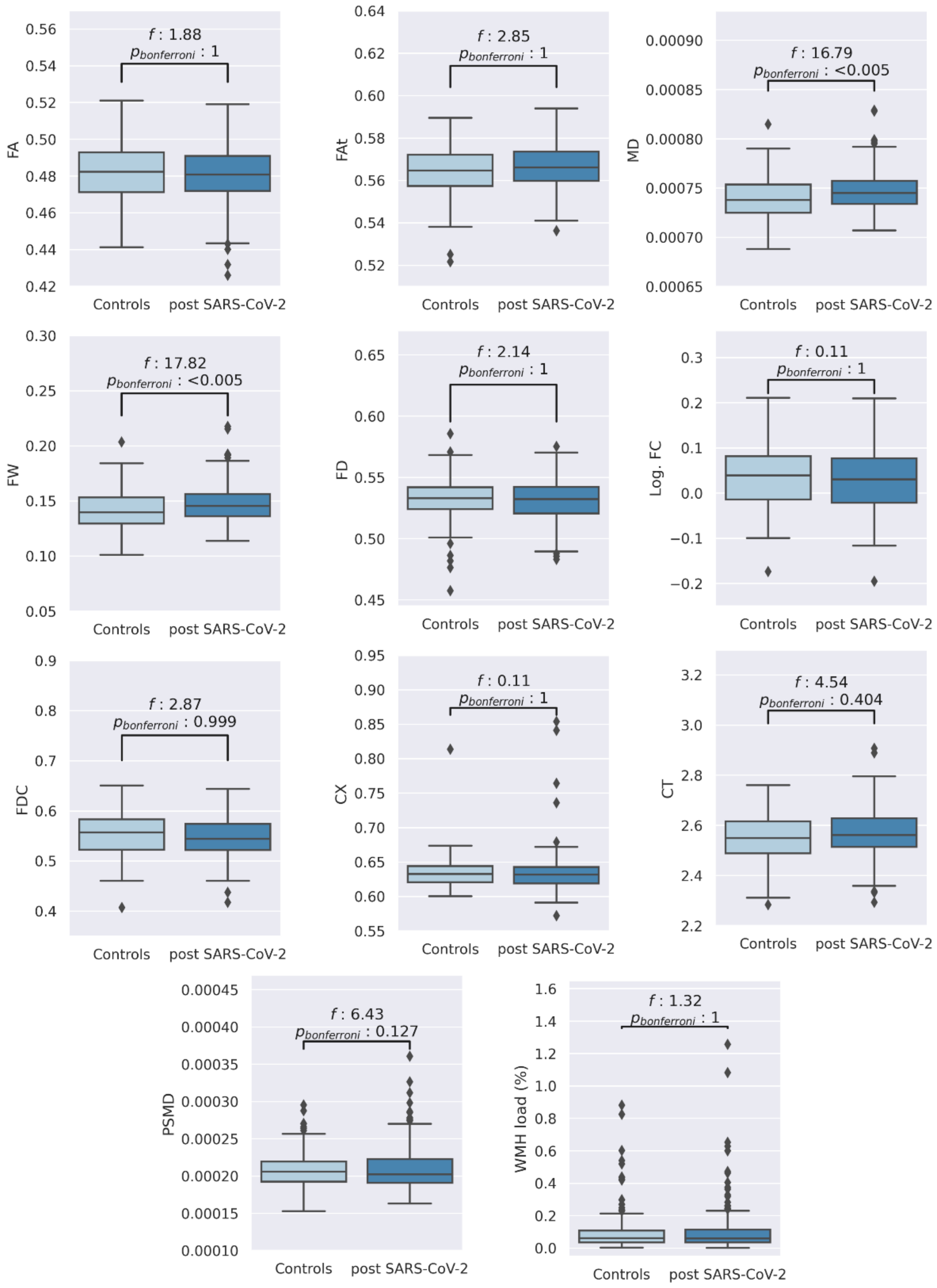

Figure S4 shows boxplots of averaged imaging measures and the corresponding statistical results (F-statistics and Bonferroni-corrected *P* values) from the ANCOVAs comparing matched controls with non-hospitalized post-SARS-CoV-2 individuals adjusted for age, sex, and years of education.

*Abbreviations*: CT = cortical thickness, CX = complexity, FA = fractional anisotropy, $FA_T$ = FA of the tissue, FD = fiber density, FDC = fiber density and cross-section, FW = free-water, Log. FC = logarithm of fiber cross-section, MD = mean diffusivity, post-SARS-CoV-2 = individuals who recovered from a severe acute respiratory coronavirus type 2 infection, PSMD = peak width of skeletonized MD, WMH = white matter hyperintensity

**Table S7. Results of clinical and neuropsychological assessments of non-hospitalized post-SARS-CoV-2 individuals compared to matched controls**

| Clinical measure[a] | Non-hospitalized Post-SARS-CoV-2 | Matched controls | $P_{uncorr}$[b] | $P_{bonf}$[c] | *F* |
|---|---|---|---|---|---|
| **Neurocognition** | | | | | |
| **TMT-A in seconds** | 31.41 ± 10.72 (194) | 33.71 ± 11.67 (190) | .07 | .62 | 3.32 |
| **TMT-B in seconds** | 67.79 ± 22.43 (194) | 70.89 ± 25.57 (187) | .27 | >.99 | 1.22 |
| **VF** | 28.00 ± 6.08 (194) | 26.43 ± 7.15 (212) | .03 | .25 | 4.90 |
| **WLR** | 8.54 ± 1.59 (193) | 8.32 ± 1.61 (204) | .27 | >.99 | 1.23 |
| **MMSE** | 28.44 ± 1.23 (193) | 28.02 ± 1.72 (210) | .009 | .08 | 6.87 |
| **CDT** | 6.76 ± 0.79 (194) | 6.57 ± 1.03 (214) | .05 | .41 | 4.03 |
| **Psychosocial symptom burden** | | | | | |
| **PHQ-9** | 3.88 ± 3.74 (195) | 3.91 ± 3.77 (215) | .83 | >.99 | 0.05 |
| **GAD-7** | 2.93 ± 3.32 (195) | 2.80 ± 3.06 (215) | .76 | >.99 | 0.09 |
| **Neurological symptom burden** | | | | | |
| **PHQ-15**[d] | 2.13 ± 1.85 (195) | 1.83 ± 1.73 (215) | .12 | >.99 | 2.39 |

*Abbreviations*: CDT = clock drawing test, GAD = General Anxiety Disorder, MMSE = Mini Mental State Examination, PHQ = Patient Health Questionnaire, post-SARS-CoV-2 individuals = individuals who recovered from a severe acute respiratory coronavirus type 2 infection, TMT-A = Trail-Making-Test Part A, TMT-B = TMT Part B, VF = verbal fluency, WLR = word list recall

[a]Presented as mean ± standard deviation (N)
[b]Uncorrected P values of analyses of covariance, adjusted for age, sex and years of education
[c]Bonferroni-corrected P values of analyses of covariance, adjusted for age, sex and years of education (considering 9 comparisons)
[d]PHQ-15 items: headache, dizziness, fatigue, sleep disturbances

**Figure S5. Machine learning prediction results excluding formerly hospitalized post-SARS-CoV-2 individuals**

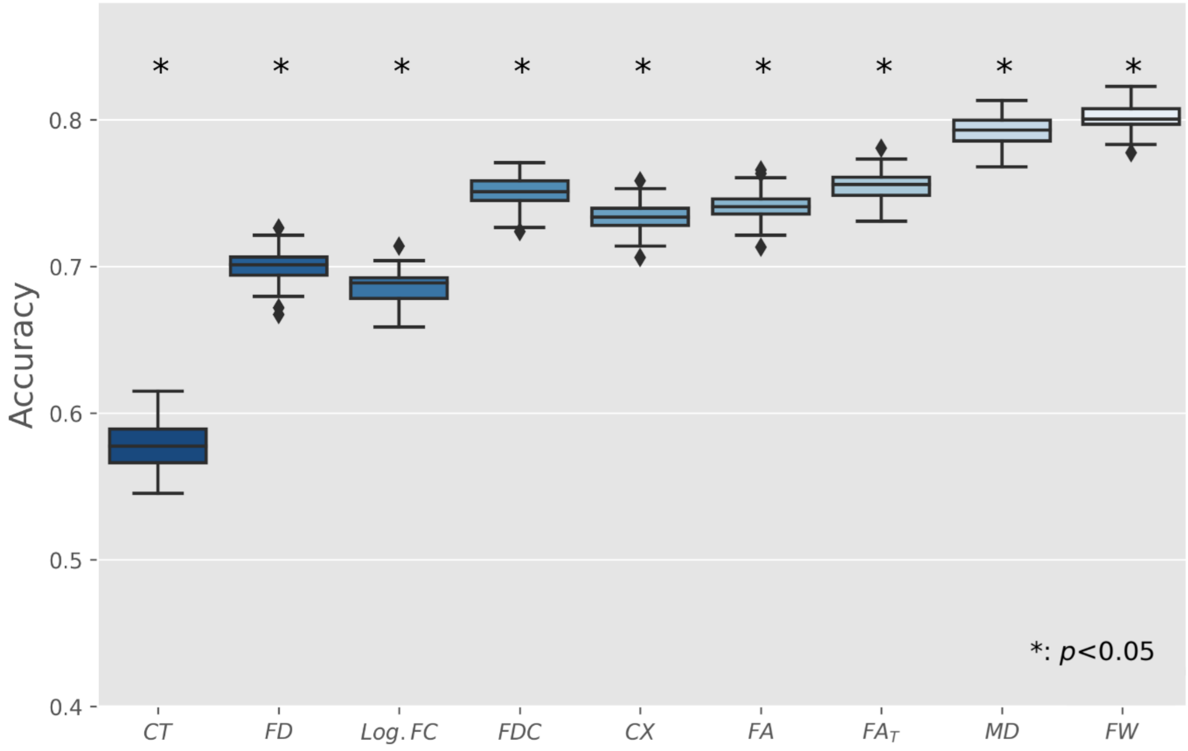

Boxplots represent the accuracy of models trained in a 10-fold nested cross-validation setup. To address scoring being biased by a single arbitrary split of training and test sets, predictions have been repeated 100 times for each marker with different random split regimens. Asterisks indicate significant difference to null-model predictions.

*Abbreviations*: CT = cortical thickness, CX = complexity, FA = fractional anisotropy, $FA_T$ = FA of the tissue, FD = fiber density, FDC = fiber density and cross-section, FW = free-water, Log. FC = logarithm of fiber cross-section, MD = mean diffusivity, post-SARS-CoV-2 = individuals who recovered from a severe acute respiratory coronavirus type 2 infection

#### Comparison of matched controls with post-SARS-CoV-2 individuals stratified by recruitment route

**Table S8. Results of analyses of covariance comparing averaged imaging markers between post-SARS-CoV-2 individuals identified via laboratory reports from our clinical information system and matched controls**

| Imaging metric[a] | Laboratory report-identified Post-SARS-CoV-2 | Matched controls | $P_{uncorr}$[b] | $P_{bonf}$[c] | *F* |
|---|---|---|---|---|---|
| **FA** | 0.478 ± 0.018 (85) | 0.482 ± 0.016 (206) | .08 | .92 | 3.02 |
| **MD ($10^{-3}$ $mm^2/s$)** | 0.751 ± 0.023 (85) | 0.740 ± 0.020 (206) | <.001 | **<.001*** ** | 18.86 |
| **$FA_T$** | 0.566 ± 0.011 (85) | 0.564 ± 0.011 (206) | .27 | >.99 | 1.21 |
| **FW** | 0.151 ± 0.020 (85) | 0.142 ± 0.017 (206) | <.001 | **<.001*** ** | 19.19 |
| **FD** | 0.527 ± 0.043 (84) | 0.531 ± 0.031 (203) | .32 | >.99 | .97 |
| **FDC** | 0.541 ± 0.068 (84) | 0.551 ± 0.055 (203) | .13 | >.99 | 2.21 |
| **Log. FC** | 0.014 ± 0.173 (84) | 0.016 ± 0.198 (205) | .89 | >.99 | .02 |
| **CX** | 0.630 ± 0.030 (84) | 0.634 ± 0.020 (203) | .22 | >.99 | 1.49 |
| **PSMD ($10^{-3}$ $mm^2/s$)** | 0.215 ± 0.032 (85) | 0.207 ± 0.023 (206) | .008 | .08 | 7.25 |
| **WMH Load (%)** | 0.110 ± 0.137 (77) | 0.099 ± 0.122 (207) | .50 | >.99 | .45 |
| **CT (mm)** | 2.561 ± 0.089 (85) | 2.550 ± 0.094 (221) | .25 | >.99 | 1.35 |

*Abbreviations*: CT = cortical thickness, CX = complexity, FA = fractional anisotropy, $FA_T$ = FA of the tissue, FD = fiber density, FDC = fiber density and cross-section, FW = free-water, Log. FC = logarithm of fiber cross-section, MD = mean diffusivity, post-SARS-CoV-2 = individuals who recovered from a severe acute respiratory coronavirus type 2 infection, PSMD = peak width of skeletonized MD, WMH = white matter hyperintensity

[a]Presented as mean ± standard deviation (N)
[b]Uncorrected *P* values of analyses of covariance, adjusted for age, sex and years of education
[c]Bonferroni-corrected *P* values of analyses of covariance, adjusted for age, sex and years of education (considering 11 comparisons)
***Denotes statistical significance at bonferroni-corrected *P* <.001

**Figure S6. Boxplots and statistics of averaged imaging markers comparing post-SARS-CoV-2 individuals identified via laboratory reports from our clinical information system with matched controls**

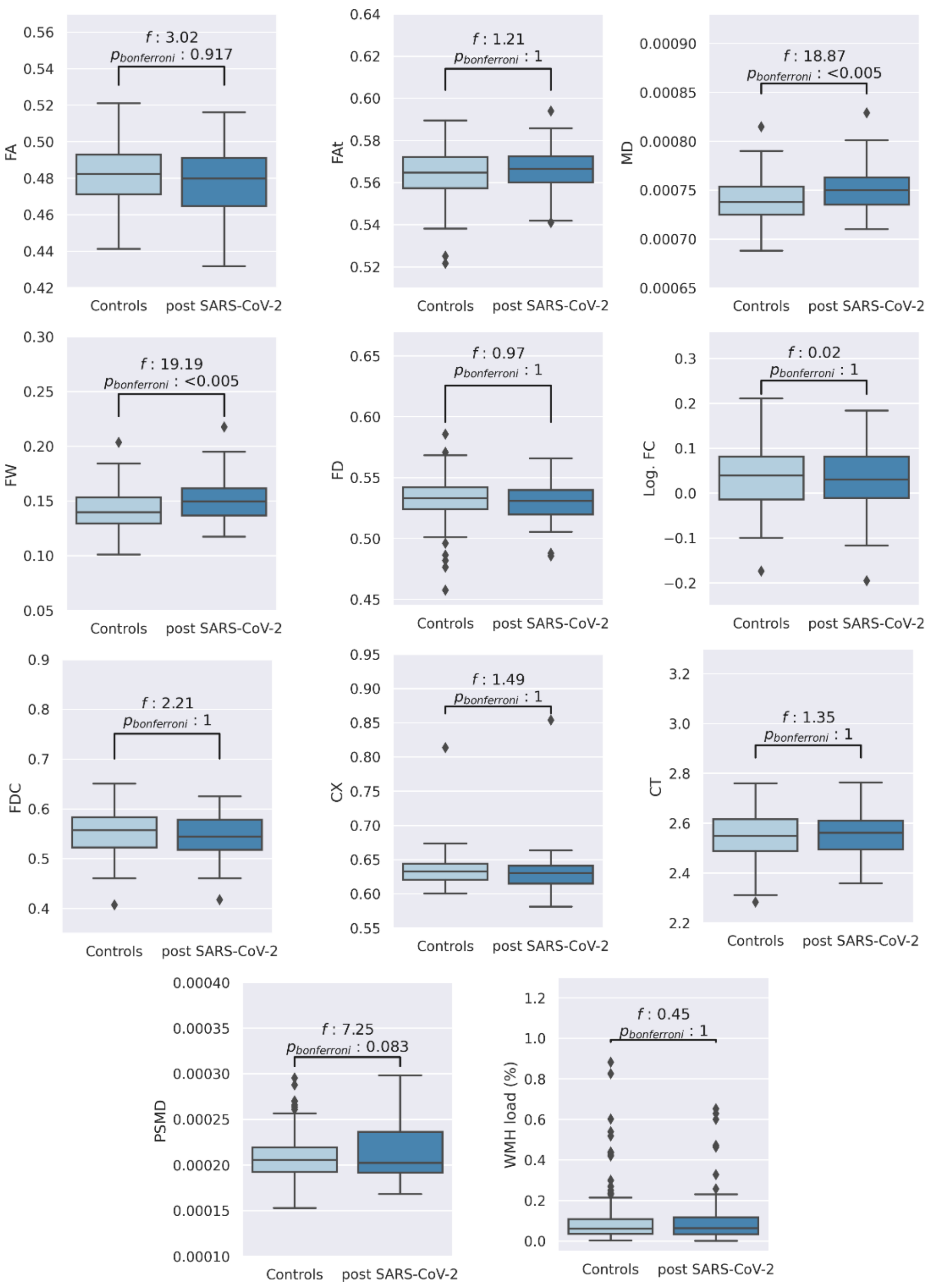

Figure S6 shows boxplots of averaged imaging measures and the corresponding statistical results (F-statistics and Bonferroni-corrected *P* values) from the ANCOVAs comparing matched controls with post-SARS-CoV-2 individuals identified via laboratory reports, adjusted for age, sex, and years of education.

*Abbreviations*: CT = cortical thickness, CX = complexity, FA = fractional anisotropy, $FA_T$ = FA of the tissue, FD = fiber density, FDC = fiber density and cross-section, FW = free-water, Log. FC = logarithm of fiber cross-section, MD = mean diffusivity, post-SARS-CoV-2 = individuals who recovered from a severe acute respiratory coronavirus type 2 infection, PSMD = peak width of skeletonized MD, WMH = white matter hyperintensity

**Table S9. Results of clinical and neuropsychological assessments of post-SARS-CoV-2 individuals identified via laboratory reports from our clinical information system compared to matched controls**

| Clinical measure[a] | Laboratory report-identified Post-SARS-CoV-2 | Matched controls | $P_{uncorr}$[b] | $P_{bonf}$[c] | *F* |
|---|---|---|---|---|---|
| **Neurocognition** | | | | | |
| **TMT-A in seconds** | 31.82 ± 9.65 (77) | 33.71 ± 11.67 (190) | .20 | >.99 | 1.65 |
| **TMT-B in seconds** | 67.13 ± 21.31 (77) | 70.89 ± 25.57 (187) | .17 | >.99 | 1.87 |
| **VF** | 27.27 ± 6.19 (77) | 26.43 ± 7.15 (212) | .30 | >.99 | 1.09 |
| **WLR** | 8.44 ± 1.79 (75) | 8.32 ± 1.61 (204) | .50 | >.99 | 0.45 |
| **MMSE** | 28.35 ± 1.23 (77) | 28.02 ± 1.72 (210) | .13 | >.99 | 2.34 |
| **CDT** | 6.75 ± 0.76 (77) | 6.57 ± 1.03 (214) | .17 | >.99 | 1.90 |
| **Psychosocial symptom burden** | | | | | |
| **PHQ-9** | 3.53 ± 3.66 (76) | 3.91 ± 3.77 (215) | .45 | >.99 | 0.56 |
| **GAD-7** | 2.49 ± 2.73 (76) | 2.80 ± 3.06 (215) | .46 | >.99 | 0.55 |
| **Neurological symptom burden** | | | | | |
| **PHQ-15**[d] | 1.91 ± 1.76 (76) | 1.83 ± 1.73 (215) | .66 | >.99 | 0.19 |

*Abbreviations*: CDT = clock drawing test, GAD = General Anxiety Disorder, MMSE = Mini Mental State Examination, PHQ = Patient Health Questionnaire, post-SARS-CoV-2 individuals = individuals who recovered from a severe acute respiratory coronavirus type 2 infection, TMT-A = Trail-Making-Test Part A, TMT-B = TMT Part B, VF = verbal fluency, WLR = word list recall

[a]Presented as mean ± standard deviation (N)
[b]Uncorrected P values of analyses of covariance, adjusted for age, sex and years of education
[c]Bonferroni-corrected P values of analyses of covariance, adjusted for age, sex and years of education (considering 9 comparisons)
[d]PHQ-15 items: headache, dizziness, fatigue, sleep disturbances

**Table S10. Results of analyses of covariance comparing averaged imaging markers between post-SARS-CoV-2 individuals identified via a newspaper announcement and matched controls**

| Imaging metric[a] | Newspaper-identified Post-SARS-CoV-2 | Matched controls | $P_{uncorr}$[b] | $P_{bonf}$[c] | *F* |
|---|---|---|---|---|---|
| **FA** | 0.481 ± 0.015 (148) | 0.482 ± 0.016 (206) | .62 | >.99 | .24 |
| **MD ($10^{-3}$ $mm^2/s$)** | 0.745 ± 0.019 (148) | 0.740 ± 0.020 (206) | .006 | .06 | 7.79 |
| **$FA_T$** | 0.566 ± 0.009 (148) | 0.564 ± 0.011 (206) | .04 | .49 | 4.07 |
| **FW** | 0.147 ± 0.016 (148) | 0.142 ± 0.017 (206) | .003 | **.035*** | 8.82 |
| **FD** | 0.525 ± 0.055 (147) | 0.531 ± 0.031 (203) | .20 | >.99 | 1.65 |
| **FDC** | 0.539 ± 0.077 (147) | 0.551 ± 0.055 (203) | .19 | >.99 | 1.74 |
| **Log. FC** | 0.006 ± 0.191 (147) | 0.016 ± 0.198 (205) | .89 | >.99 | .02 |
| **CX** | 0.634 ± 0.029 (147) | 0.634 ± 0.020 (203) | .70 | >.99 | .15 |
| **PSMD ($10^{-3}$ $mm^2/s$)** | 0.210 ± 0.030 (148) | 0.207 ± 0.023 (206) | .045 | .50 | 4.04 |
| **WMH Load (%)** | 0.101 ± 0.152 (148) | 0.099 ± 0.122 (207) | .65 | >.99 | .20 |
| **CT (mm)** | 2.577 ± 0.010 (139) | 2.550 ± 0.094 (221) | .008 | .09 | 7.17 |

*Abbreviations*: CT = cortical thickness, CX = complexity, FA = fractional anisotropy, $FA_T$ = FA of the tissue, FD = fiber density, FDC = fiber density and cross-section, FW = free-water, Log. FC = logarithm of fiber cross-section, MD = mean diffusivity, post-SARS-CoV-2 = individuals who recovered from a severe acute respiratory coronavirus type 2 infection, PSMD = peak width of skeletonized MD, WMH = white matter hyperintensity

[a]Presented as mean ± standard deviation (N)
[b]Uncorrected *P* values of analyses of covariance, adjusted for age, sex and years of education
[c]Bonferroni-corrected *P* values of analyses of covariance, adjusted for age, sex and years of education (considering 11 comparisons)
*Denotes statistical significance at bonferroni-corrected *P* <.05

**Figure S7. Boxplots and statistics of averaged imaging markers comparing post-SARS-CoV-2 individuals identified via a newspaper announcement with matched controls**

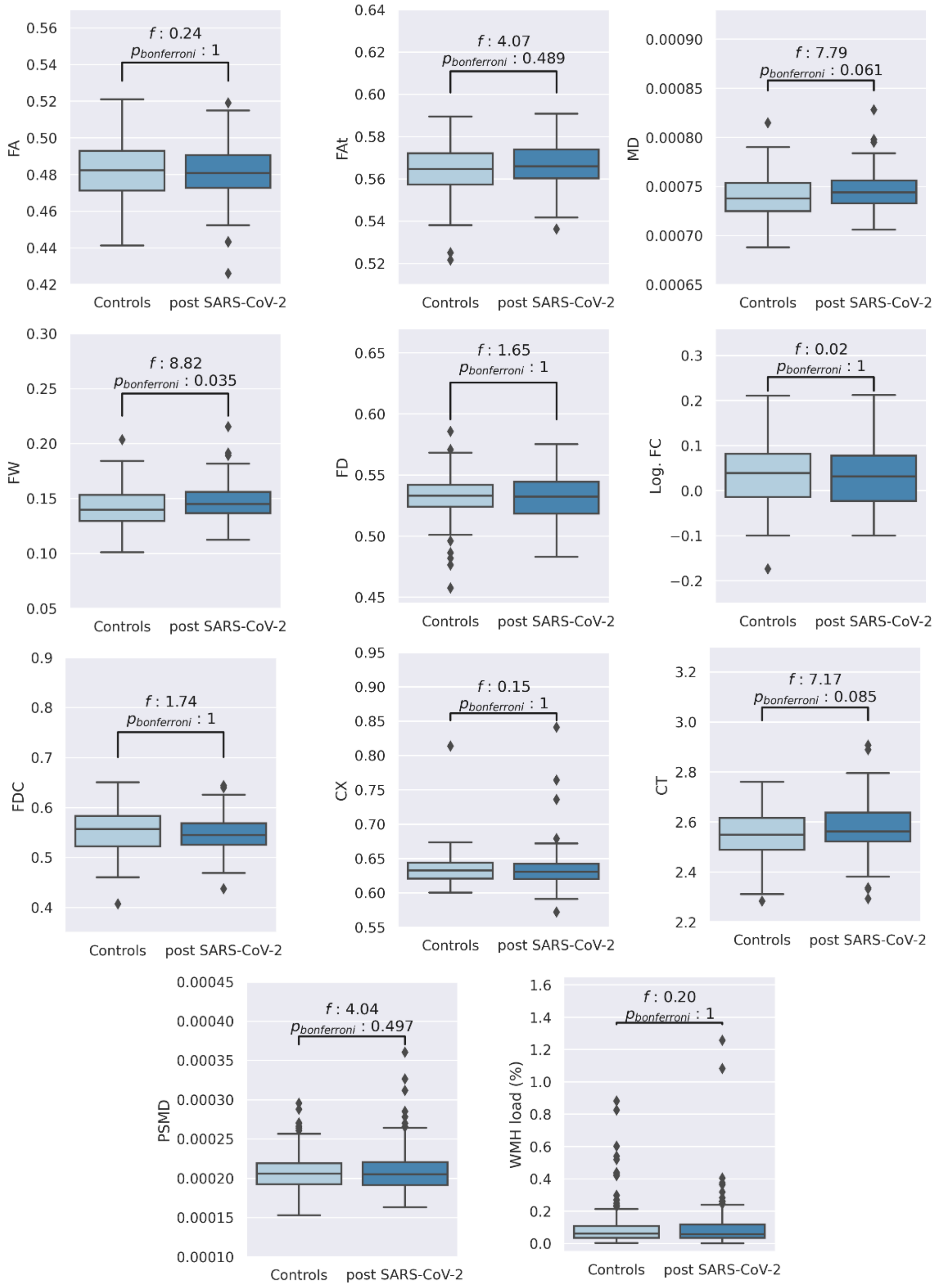

Figure S7 shows boxplots of averaged imaging measures and the corresponding statistical results (F-statistics and Bonferroni-corrected *P* values) from the ANCOVAs comparing matched controls with post-SARS-CoV-2 individuals identified via newspaper announcement, adjusted for age, sex, and years of education.

*Abbreviations*: CT = cortical thickness, CX = complexity, FA = fractional anisotropy, $FA_T$ = FA of the tissue, FD = fiber density, FDC = fiber density and cross-section, FW = free-water, Log. FC = logarithm of fiber cross-section, MD = mean diffusivity, post-SARS-CoV-2 = individuals who recovered from a severe acute respiratory coronavirus type 2 infection, PSMD = peak width of skeletonized MD, WMH = white matter hyperintensity

**Table S11. Results of clinical and neuropsychological assessments of post-SARS-CoV-2 individuals identified via a newspaper announcement compared to matched controls**

| Clinical measure[a] | Newspaper-identified Post-SARS-CoV-2 | Matched controls | $P_{uncorr}^{b}$ | $P_{bonf}^{c}$ | *F* |
|---|---|---|---|---|---|
| **Neurocognition** | | | | | |
| **TMT-A in seconds** | 31.93 ± 11.13 (135) | 33.71 ± 11.67 (190) | .22 | >.99 | 1.52 |
| **TMT-B in seconds** | 69.29 ± 23.49 (135) | 70.89 ± 25.57 (187) | .75 | >.99 | 0.10 |
| **VF** | 28.46 ± 5.94 (135) | 26.43 ± 7.15 (212) | .01 | .10 | 6.45 |
| **WLR** | 8.56 ± 1.55 (135) | 8.32 ± 1.61 (204) | .27 | >.99 | 1.21 |
| **MMSE** | 28.39 ± 1.29 (134) | 28.02 ± 1.72 (210) | .05 | .43 | 3.95 |
| **CDT** | 6.76 ± 0.79 (135) | 6.57 ± 1.03 (214) | .07 | .65 | 3.25 |
| **Psychosocial symptom burden** | | | | | |
| **PHQ-9** | 4.18 ± 3.77 (136) | 3.91 ± 3.77 (215) | .60 | >.99 | 0.28 |
| **GAD-7** | 3.20 ± 3.53 (136) | 2.80 ± 3.06 (215) | .31 | >.99 | 1.04 |
| **Neurological symptom burden** | | | | | |
| **PHQ-15[d]** | 2.25 ± 1.86 (136) | 1.83 ± 1.73 (215) | .04 | .40 | 4.07 |

*Abbreviations*: CDT = clock drawing test, GAD = General Anxiety Disorder, MMSE = Mini Mental State Examination, PHQ = Patient Health Questionnaire, post-SARS-CoV-2 individuals = individuals who recovered from a severe acute respiratory coronavirus type 2 infection, TMT-A = Trail-Making-Test Part A, TMT-B = TMT Part B, VF = verbal fluency, WLR = word list recall

[a]Presented as mean ± standard deviation (N)
[b]Uncorrected P values of analyses of covariance, adjusted for age, sex and years of education
[c]Bonferroni-corrected P values of analyses of covariance, adjusted for age, sex and years of education (considering 9 comparisons)
[d]PHQ-15 items: headache, dizziness, fatigue, sleep disturbances

#### Code availability

**Table S9. URLs to GitHub repositories containing analysis code**

| **Analysis step** | ***Code URL*** |
|---|---|
| DWI preprocessing with QSIPrep | https://github.com/csi-hamburg/CSIframe/blob/709275c816b7746bf7168f69b652b2aec569b838/pipelines/qsiprep/qsiprep.sh |
| Fixel-based analysis | https://github.com/csi-hamburg/CSIframe/tree/main/pipelines/fba |
| Free-water and diffusion tensor imaging | https://github.com/csi-hamburg/CSIframe/blob/main/pipelines/freewater/freewater.sh |
| Peak-width of skeletonized mean diffusivity | https://github.com/csi-hamburg/CSIframe/blob/main/pipelines/psmd/psmd_csi.sh |
| Statistics | https://github.com/csi-hamburg/2022_petersen_naegele_postcovid_imaging |
| Structural processing with CAT | https://github.com/csi-hamburg/CSIframe/blob/709275c816b7746bf7168f69b652b2aec569b838/pipelines/cat12/cat12.sh |
| Tract-based spatial statistics | https://github.com/csi-hamburg/CSIframe/tree/main/pipelines/tbss |
| Voxel-wise statistics of diffusion markers | https://github.com/csi-hamburg/CSIframe/tree/main/pipelines/statistics |
| White matter hyperintensity segmentation | https://github.com/csi-hamburg/CSIframe/tree/main/pipelines/wmh |

#### References

1. Cieslak M, Cook PA, He X, et al. QSIPrep: an integrative platform for preprocessing and reconstructing diffusion MRI data. *Nat Methods*. 2021;18(7):775-778. doi:10.1038/s41592-021-01185-5

2. Gorgolewski K, Burns C, Madison C, et al. Nipype: A Flexible, Lightweight and Extensible Neuroimaging Data Processing Framework in Python. *Frontiers in Neuroinformatics*. 2011;5. Accessed March 28, 2022. https://www.frontiersin.org/article/10.3389/fninf.2011.00013

3. Tustison NJ, Avants BB, Cook PA, et al. N4ITK: improved N3 bias correction. *IEEE Trans Med Imaging*. 2010;29(6):1310-1320. doi:10.1109/TMI.2010.2046908

4. Zhang Y, Brady M, Smith S. Segmentation of brain MR images through a hidden Markov random field model and the expectation-maximization algorithm. *IEEE Trans Med Imaging*. 2001;20(1):45-57. doi:10.1109/42.906424

5. Gaser C, Dahnke R. CAT-a computational anatomy toolbox for the analysis of structural MRI data. *Hbm*. 2016;2016:336-348.

6. Dahnke R, Yotter RA, Gaser C. Cortical thickness and central surface estimation. *NeuroImage*. 2013;65:336-348. doi:10.1016/j.neuroimage.2012.09.050

7. Yotter RA, Dahnke R, Thompson PM, Gaser C. Topological correction of brain surface meshes using spherical harmonics. *Hum Brain Mapp*. 2011;32(7):1109-1124. doi:10.1002/hbm.21095

8. Yotter RA, Thompson PM, Gaser C. Algorithms to Improve the Reparameterization of Spherical Mappings of Brain Surface Meshes. *Journal of Neuroimaging*. 2011;21(2):e134-e147. doi:10.1111/j.1552-6569.2010.00484.x

9. Veraart J, Novikov DS, Christiaens D, Ades-aron B, Sijbers J, Fieremans E. Denoising of diffusion MRI using random matrix theory. *Neuroimage*. 2016;142:394-406. doi:10.1016/j.neuroimage.2016.08.016

10. Kellner E, Dhital B, Kiselev VG, Reisert M. Gibbs-ringing artifact removal based on local subvoxel-shifts. *Magn Reson Med*. 2016;76(5):1574-1581. doi:10.1002/mrm.26054

11. Andersson JLR, Sotiropoulos SN. An integrated approach to correction for off-resonance effects and subject movement in diffusion MR imaging. *Neuroimage*. 2016;125:1063-1078. doi:10.1016/j.neuroimage.2015.10.019

12. Esteban O, Markiewicz CJ, Blair RW, et al. fMRIPrep: a robust preprocessing pipeline for functional MRI. *Nat Methods*. 2019;16(1):111-116. doi:10.1038/s41592-018-0235-4

13. Huntenburg JM, Gorgolewski KJ, Anwander A, Margulies D. Evaluating nonlinear coregistration of BOLD EPI and T1 images. Published online 2014. doi:10.7490/F1000RESEARCH.1096036.1

14. Power JD, Mitra A, Laumann TO, Snyder AZ, Schlaggar BL, Petersen SE. Methods to detect, characterize, and remove motion artifact in resting state fMRI. *Neuroimage*. 2014;84:320-341. doi:10.1016/j.neuroimage.2013.08.048

15. Abraham A, Pedregosa F, Eickenberg M, et al. Machine learning for neuroimaging with scikit-learn. *Frontiers in Neuroinformatics*. 2014;8. Accessed March 28, 2022. https://www.frontiersin.org/article/10.3389/fninf.2014.00014

16. Garyfallidis E, Brett M, Amirbekian B, et al. Dipy, a library for the analysis of diffusion MRI data. *Frontiers in Neuroinformatics*. 2014;8. Accessed March 28, 2022. https://www.frontiersin.org/article/10.3389/fninf.2014.00008

17. Basser PJ, Pierpaoli C. Microstructural and physiological features of tissues elucidated by quantitative-diffusion-tensor MRI. *J Magn Reson B*. 1996;111(3):209-219. doi:10.1006/jmrb.1996.0086

18. Basser PJ, Mattiello J, LeBihan D. MR diffusion tensor spectroscopy and imaging. *Biophys J*. 1994;66(1):259-267. doi:10.1016/S0006-3495(94)80775-1

19. Pasternak O, Sochen N, Gur Y, Intrator N, Assaf Y. Free water elimination and mapping from diffusion MRI. *Magnetic Resonance in Medicine*. 2009;62(3):717-730. doi:10.1002/mrm.22055

20. Tournier JD, Smith R, Raffelt D, et al. MRtrix3: A fast, flexible and open software framework for medical image processing and visualisation. *NeuroImage*. 2019;202:116137. doi:10.1016/j.neuroimage.2019.116137

21. Dhollander T, Mito R, Raffelt D, Connelly A. Improved white matter response function estimation for 3-tissue constrained spherical deconvolution. Published online 2019:10.

22. Tournier JD, Yeh CH, Calamante F, Cho KH, Connelly A, Lin CP. Resolving crossing fibres using constrained spherical deconvolution: validation using diffusion-weighted imaging phantom data. *Neuroimage*. 2008;42(2):617-625. doi:10.1016/j.neuroimage.2008.05.002

23. Dhollander T, Connelly A. A novel iterative approach to reap the benefits of multi-tissue CSD from just single-shell (+b=0) diffusion MRI data. Published online 2016:8.

24. Dhollander T, Mito R, Raffelt D, Connelly A. Improved white matter response function estimation for 3-tissue constrained spherical deconvolution. Published online 2019:10.

25. (ISMRM 2017) Bias Field Correction and Intensity Normalisation for Quantitative Analysis of Apparent Fibre Density. Accessed March 28, 2022. https://ismrm.gitlab.io/2017/3541.html

26. Tournier J, Calamante F, Connelly A. Improved probabilistic streamlines tractography by 2 nd order integration over fibre orientation distributions. Published 2009. Accessed April 5, 2022. https://www.semanticscholar.org/paper/Improved-probabilistic-streamlines-tractography-by-Tournier-Calamante/b4ffcb9ec889a8a68bffc46387a96b78a50ef94a

27. Smith RE, Tournier JD, Calamante F, Connelly A. SIFT: Spherical-deconvolution informed filtering of tractograms. *Neuroimage*. 2013;67:298-312. doi:10.1016/j.neuroimage.2012.11.049

28. Wasserthal J, Neher P, Maier-Hein KH. TractSeg - Fast and accurate white matter tract segmentation. *NeuroImage*. 2018;183:239-253. doi:10.1016/j.neuroimage.2018.07.070

29. Raffelt DA, Tournier JD, Smith RE, et al. Investigating white matter fibre density and morphology using fixel-based analysis. *Neuroimage*. 2017;144(Pt A):58-73. doi:10.1016/j.neuroimage.2016.09.029

30. Riffert TW, Schreiber J, Anwander A, Knösche TR. Beyond fractional anisotropy: extraction of bundle-specific structural metrics from crossing fiber models. *Neuroimage*. 2014;100:176-191. doi:10.1016/j.neuroimage.2014.06.015

31. Avants BB, Epstein CL, Grossman M, Gee JC. Symmetric diffeomorphic image registration with cross-correlation: evaluating automated labeling of elderly and neurodegenerative brain. *Med Image Anal*. 2008;12(1):26-41. doi:10.1016/j.media.2007.06.004

32. Smith SM, Jenkinson M, Johansen-Berg H, et al. Tract-based spatial statistics: Voxelwise analysis of multi-subject diffusion data. *NeuroImage*. 2006;31(4):1487-1505. doi:10.1016/j.neuroimage.2006.02.024

33. Jenkinson M, Beckmann CF, Behrens TEJ, Woolrich MW, Smith SM. FSL. *Neuroimage*. 2012;62(2):782-790. doi:10.1016/j.neuroimage.2011.09.015

34. Baykara E, Gesierich B, Adam R, et al. A Novel Imaging Marker for Small Vessel Disease Based on Skeletonization of White Matter Tracts and Diffusion Histograms. *Annals of Neurology*. 2016;80(4):581-592. doi:10.1002/ana.24758

35. Griffanti L, Zamboni G, Khan A, et al. BIANCA (Brain Intensity AbNormality Classification Algorithm): A new tool for automated segmentation of white matter hyperintensities. *Neuroimage*. 2016;141:191-205. doi:10.1016/j.neuroimage.2016.07.018

36. Sundaresan V, Zamboni G, Le Heron C, et al. Automated lesion segmentation with BIANCA: Impact of population-level features, classification algorithm and locally adaptive thresholding. *NeuroImage*. 2019;202:116056. doi:10.1016/j.neuroimage.2019.116056

37. Jenkinson M, Smith S. A global optimisation method for robust affine registration of brain images. *Med Image Anal*. 2001;5(2):143-156. doi:10.1016/s1361-8415(01)00036-6

38. Jenkinson M, Bannister P, Brady M, Smith S. Improved optimization for the robust and accurate linear registration and motion correction of brain images. *Neuroimage*. 2002;17(2):825-841. doi:10.1016/s1053-8119(02)91132-8

39. Avants B, Tustison NJ, Song G. Advanced Normalization Tools: V1.0. *The Insight Journal*. Published online July 29, 2009. doi:10.54294/uvnhin

40. Fischl B, Salat DH, Busa E, et al. Whole brain segmentation: automated labeling of neuroanatomical structures in the human brain. *Neuron*. 2002;33(3):341-355. doi:10.1016/s0896-6273(02)00569-x

41. Esteban O, Birman D, Schaer M, Koyejo OO, Poldrack RA, Gorgolewski KJ. MRIQC: Advancing the automatic prediction of image quality in MRI from unseen sites. Bernhardt BC, ed. *PLoS ONE*. 2017;12(9):e0184661. doi:10.1371/journal.pone.0184661

42. Fischl B, Dale AM. Measuring the thickness of the human cerebral cortex from magnetic resonance images. *Proceedings of the National Academy of Sciences*. 2000;97(20):11050-11055. doi:10.1073/pnas.200033797

43. Desikan RS, Ségonne F, Fischl B, et al. An automated labeling system for subdividing the human cerebral cortex on MRI scans into gyral based regions of interest. *Neuroimage*. 2006;31(3):968-980. doi:10.1016/j.neuroimage.2006.01.021

44. Zou H, Hastie T. Regularization and variable selection via the elastic net. *Journal of the Royal Statistical Society: Series B (Statistical Methodology)*. 2005;67(2):301-320. doi:10.1111/j.1467-9868.2005.00503.x

45. Defazio A, Bach F, Lacoste-Julien S. SAGA: A Fast Incremental Gradient Method With Support for Non-Strongly Convex Composite Objectives. *arXiv:14070202 [cs, math, stat]*. Published online December 16, 2014. Accessed April 21, 2022. http://arxiv.org/abs/1407.0202

46. Varoquaux G, Raamana PR, Engemann DA, Hoyos-Idrobo A, Schwartz Y, Thirion B. Assessing and tuning brain decoders: Cross-validation, caveats, and guidelines. *Neuroimage*. 2017;145(Pt B):166-179. doi:10.1016/j.neuroimage.2016.10.038
